# Stochastic Morphodynamics of the Human Aorta Across the Lifespan

**DOI:** 10.64898/2026.06.05.26355015

**Authors:** Kelly C. Twohig, Michael Mansour, Joseph A. Pugar, Karen Yuan, Nhung Nguyen, Luka Pocivavsek, Andrei A. Klishin

**Affiliations:** Department of Surgery, University of Chicago, Chicago, IL; Department of Medical Physics, University of Chicago, IL; Department of Surgery, Northwestern University, Chicago, IL; Department of Mechanical Engineering, University of Hawai‘ i at Mānoa, Honolulu, HI

## Abstract

Biological systems evolve as continuous dynamical processes, but at organ-scale and across human lifespans they are rarely observed longitudinally—–population data typically exist instead as sparse, cross-sectional snapshots. Inferring lifespan dynamics from such data requires methods distinct from those used at cellular and tissue scales where dense observations are accessible. We address this problem in the thoracic aorta, where surgical decisions currently rest on static, age- and sex-agnostic diameter thresholds that reduce three-dimensional morphology to a single scalar. Treating normal aortic morphology as a stochastic dynamical system, we pose a continuous-time drift-diffusion process in a two-coordinate state space of normalized surface area (*Ã*) and normalized fluctuation in integrated Gaussian curvature 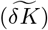, and fit closed-form solutions of the Fokker-Planck equation by maximum likelihood to a sex-balanced, age-uniform cohort spanning infancy to age 99. Inter-individual variability is treated as a fitted diffusion parameter rather than as residual scatter, which is distinct from prior normative studies that report variability as scatter around a regression line. The framework identifies two growth regimes for aortic size (childhood expansion followed by persistent adult growth, with adult males growing approximately 70% faster than adult females) and a single dynamical regime for aortic shape, with heteroscedastic variability accumulating at a rate comparable to the mean drift over the lifespan. Applied to independent cohorts of acute and chronic thoracic aortic dissections, the multivariate model identifies over 95% as statistical outliers via Mahalanobis distance, consistently outperforming either coordinate alone. The same probabilistic envelope that describes normal aging thus defines a baseline against which disease can be detected, supporting a shift toward dynamic, age- and sex-aware assessment of thoracic aortic pathology.

## I. INTRODUCTION

Biological systems evolve as continuous dynamical processes, yet data-driven inference of governing equations has matured primarily where observations are dense, controlled, and accessible at experimental cadence. The methods for learning from such *longitudinal* data were first developed for physical science and engineering [1– before expanding to biology [7]. At cellular scales, machine-learning frameworks have inferred interpretable models of cytoskeletal mechanics [8] and cellular hydro-dynamics [9], and stochastic-process inference has been used to separate intrinsic phenotypic variability from motion stochasticity and measurement noise in single-cell trajectories [10]. At tissue scales, generative models recover multicellular dynamics from high-throughput live imaging [11], and geometric analyses capture organ-level deformation and morphogenesis from time-lapsed recordings of small animal models [12, 13]. Across these scales, dynamical inference is enabled by dense observations taken under experimental control over periods of minutes to days. Extending this approach to organ-scale data across the human lifespan is structurally harder, as individuals are rarely observed for decades, and population data exist instead as sparse, *cross-sectional* snapshots. Inferring lifespan dynamics from such data requires methods that extract population-level structure from collections of single-timepoint observations, a regime distinct from dense, longitudinal inference at smaller length scales; the focus of the present paper is precisely building such methods for human data.

The aorta is a representative case in human anatomy; as the largest blood vessel in the body, it carries blood from the heart and distributes it through branches to the main arteries of organs and limbs. In children, the aorta grows with body size [14, 15]; in healthy adults, it undergoes slow changes in both size and shape throughout life, with slight differences across biological sex [16–19]. Aortic diseases (ADs) are clinically defined by changes in aortic anatomy (morphology); in patients with aortic disease, significant increases in aortic size characterize the pathology (e.g., aneurysms, dissections) (see Fig. 1) [20–22]. The prevalence of aortic pathologies in patients over 65 is 3–5%, which is comparable to heart failure and makes it one of the most prevalent cardiovascular diseases [23, 24]. These changes in morphology can be observed and quantified through computed tomography angiography (CTA)—the primary imaging modality for diagnosing aortic disease—which provides a detailed 3D shape of an individual aorta at one moment in time [25, 26]. Applying geometric descriptors to clinical decision-making, however, requires a normative baseline from which pathologic deviations can be identified, to quantify what significant increases are. Even then, returning to a “new normal” morphology is a moving target, so the baseline itself must be a dynamical object, since what counts as “normal” at age 30 is not what counts as “normal” at age 70 [20, 27–35].

**FIG. 1.**
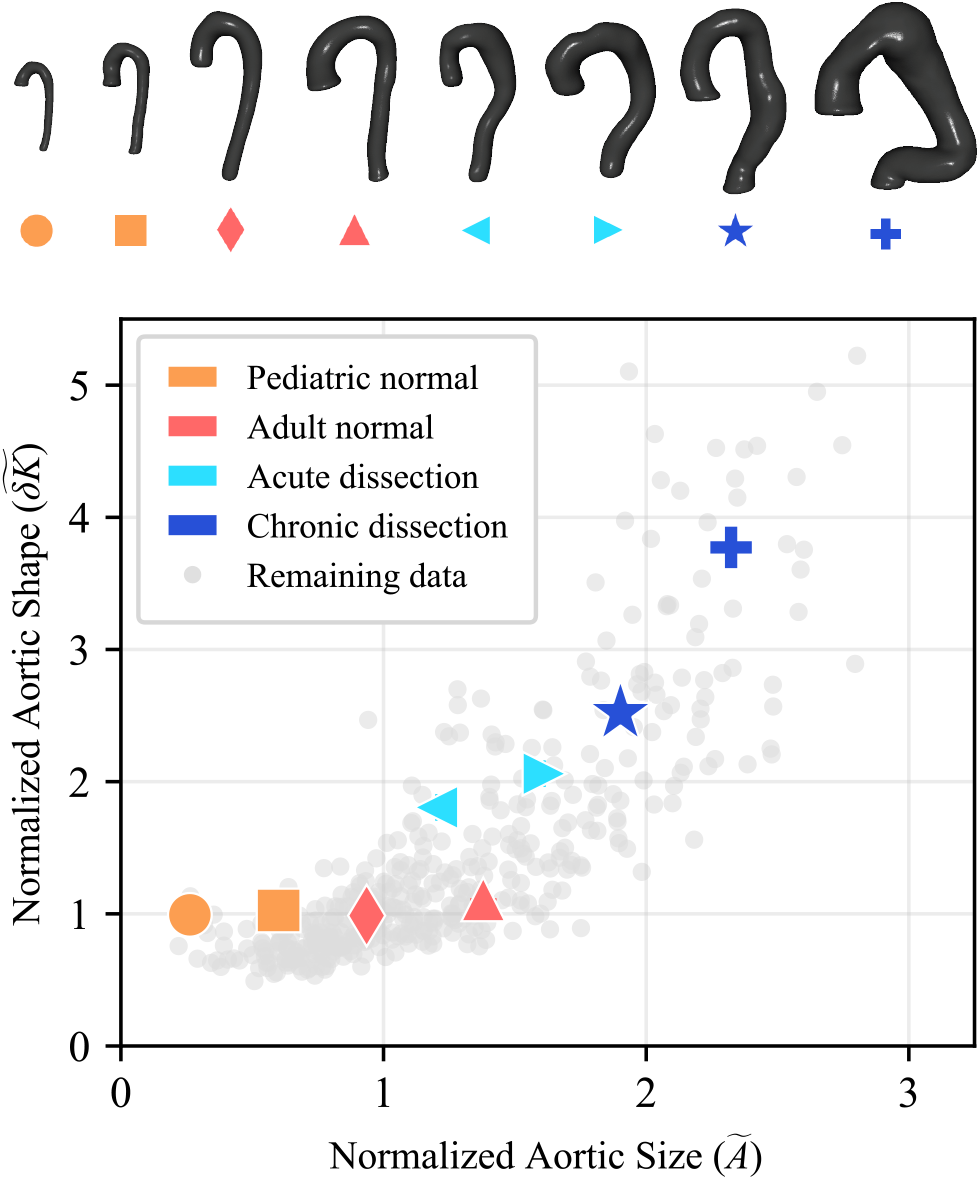
Representative segmented thoracic aortas (top) and their locations in normalized aortic size-shape space (bottom). In health, aortas undergo shape-preserving growth from childhood through adulthood. However, disease processes lead to growth and shape change, indicated by the elevated 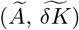 coordinates of patients with acute and chronic ADs.

For the past several decades, the primary geometric descriptor of aortic size has been the maximum aortic diameter, with thresholds for surgical repair established by landmark trials in the 1990s–2000s [36–42]. Diameter-based threshold rules nonetheless demonstrate three limitations: they do not account for inter-individual variability in normal aortic morphology, do not account for biological aging, and do not incorporate shape information available from CTA. Prior normative studies of aortic morphology [16–19] address the first two limitations only partially, reporting variability as residual scatter around piecewise-linear regression fits within discrete age bins rather than as a feature of the underlying dynamics. The third limitation of shape information is partially addressed in our recent work which supplemented diameter with a shape metric (the fluctuation in integrated Gaussian curvature, *δK*) that improved discrimination of post-repair outcomes [20], but can equally be applied to normal aortas.

In this study, we move beyond all three limitations by treating the normal aorta as a stochastic dynamical system and establishing a quantitative framework for describing its morphologic evolution across the lifespan. We curate a sex-balanced and age-uniform CTA dataset of healthy aortas with approximately equal representation in each decade from infancy to age 99, and we pose a continuous-time drift-diffusion *morphodynamic* process in a two-coordinate state space of normalized surface area (*Ã*) and normalized fluctuation in integrated Gaussian curvature 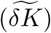. The deterministic drift *β* captures the population-mean trajectory, and the diffusion *α* captures the rate at which inter-individual variability accumulates with age. The diffusion parameter *α* is treated here as a fitted dynamical quantity rather than as a residual statistic, which distinguishes this framework from prior cross-sectional approaches that report variability as scatter around a regression line [16–19]. The approach is parsimonious, with four parameters per morphological coordinate [43, 44], and admits exact solutions to the Fokker-Planck equation [45] from which maximum-likelihood inference proceeds. Deviations from the resulting probabilistic envelope are quantified via Mahalanobis distance [46], enabling identification of morphologic outliers consistent with disease.

This work addresses the cross-sectional regime of organ-scale morphodynamics and thus more broadly learning stochastic dynamical equations from challenging cross-sectional datasets. This approach is complementary to longitudinal study of morphodynamics, in which individual trajectories are inferred from repeated scans of the same patient [47], inspired by the data-driven approaches to equation discovery in physical systems [4, 48, 49]. While this perspective focuses on different clinical questions (normal aortic evolution as opposed to post-operative recovery), it shares the underlying view of organ-scale geometry as a low-dimensional dynamical system. The paper is organized as follows: Section II describes data curation, dynamical model fitting, and outlier detection methods; Section III presents the discovered model and outlier detection results; Section IV situates other results within the literature on “normal” aortas; Section V concludes the study.

## II. METHODS

### A. Data curation

Our study uses two types of clinical patient data sets: the normal cohort for model fitting and the pathologic cohorts for statistical outlier detection. The normal cohort includes 311 patients (137 female, 174 male) with approximately equal representation in each decade from 0–9 to 90–99 years (Table IV). The cohort is predominantly Black (78.8% female, 64.9% male), reflecting the population served by the University of Chicago trauma centers. Other races include white (12.4% female, 19.5% male), more than one race (5.1% female, 8.6% male), Pacific Islander (0% female, 0.6% male), or unknown/not reported (3.7% female, 6.4% male). Where multiple CTAs were available for the same patient (77 patients: 20 female, 57 male), follow-up scans were reserved exclusively for longitudinal validation of the fitted models (Section III A 3); model fitting used only first scans. Trajectories with a between-scan axial-resolution difference greater than 1 mm were excluded from longitudinal analysis to minimize resolution-dependent segmentation error.

The pathologic cohorts are comprised of 156 pre-operative scans: 86 acute dissections (23 female, 63 male) and 70 chronic dissections (35 female, 35 male). Dissection chronicity was defined by the Society for Vascular Surgery and Society of Thoracic Surgery guidelines, where acute corresponds to an observation within 14 days of initial diagnosis, and chronic corresponds to an observation more than 90 days from initial diagnosis [50].

### B. Segmentation and morphological parameters

Three-dimensional models of the thoracic aorta were generated from CTA scans using commercial software (Simpleware; Synopsys, Inc., Sunnyvale, USA). Aortic segmentations were performed semi-automatically with PRAEVAorta Web (Nurea, Bègles, France), extending from the right coronary artery proximally to the celiac trunk distally.

To minimize surface noise prior to analysis, the segmentation masks were dilated and smoothed. Masks were then converted into two-dimensional surface meshes using Simpleware’s default meshing algorithm. Surface area and local estimations of Gaussian curvature (*K*) were calculated, and subsequently the total surface area and the fluctuation of Gaussian curvature integrated across all partitions was determined. Surface area (*A*) and *δK* were scaled by the average of the entire non-pathologic cohort (*Ā* = 243.2 cm^2^, 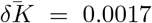) to yield normalized morphologic descriptors of aortic size (*Ã*) and shape 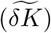. For each metric, error may be quantified as the variation due to a choice of parameterizations for meshing and partitioning the segmented aorta. This methodology has been validated and described in detail in our previous work [20, 51]. Following the methodology, each CTA scan is converted into a pair (*t*, **x**) where *t* is the chronological age of the patient at scan time and **x** is a vector of morphological parameters, in our case 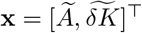.

### C. Stochastic dynamical modeling

We model the dynamics of normal aortic morphology as a stochastic system consisting of constant deterministic growth and fluctuations, decoupled across the morphological parameters. For a defined morphologic parameter *x*, the dynamics can be described by an Itô stochastic differential equation (SDE) of the following form:

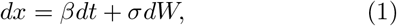

where *β* is the deterministic drift coefficient, *σ* is the diffusion coefficient, and *dW* is an increment of standard Wiener process (Brownian motion). While this SDE allows us to simulate individual stochastic growth trajectories (Fig. 2), to describe the evolution of the population of normal aortic morphology we must recast the dynamics into a probabilistic form. For a distribution *p*(*x, t*) that is subject to the same constant drift and diffusion as Eqn. 1, evolution is described by the Fokker-Planck equation [45]:

**FIG. 2.**
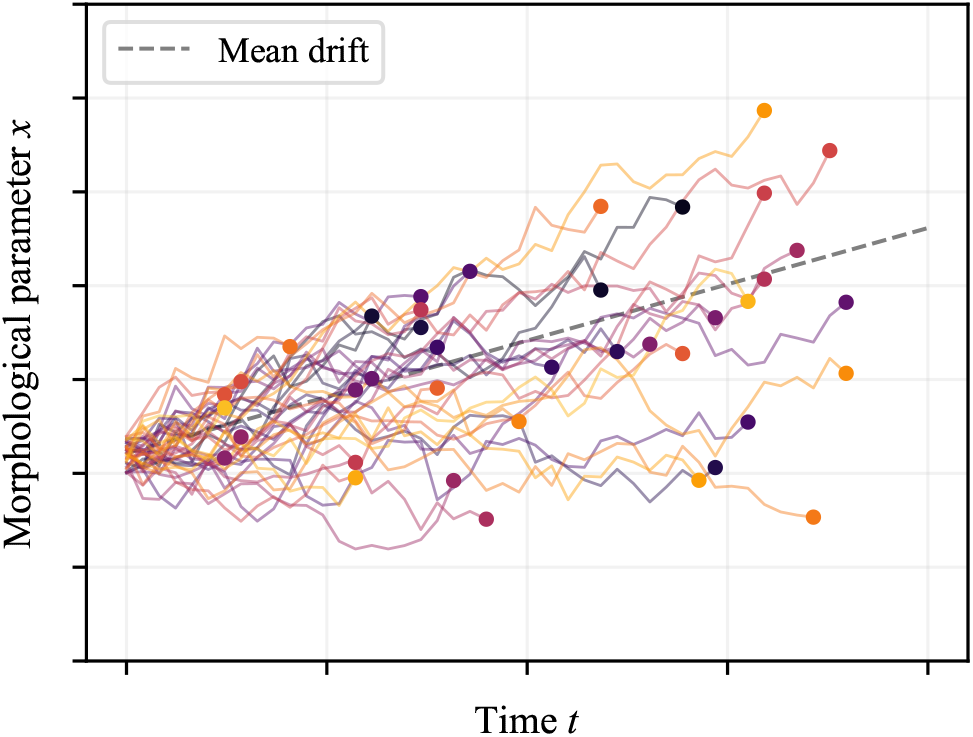
Simulated trajectories of stochastic drift-diffusion behavior (Eqn. 1). Individual trajectories evolve from a distribution of initial morphologic values under constant drift *β* and diffusion *σ*. Each dot marks the endpoint of a trajectory observed at a different time, analogous to the cross-sectional sampling of patient aortas at varying ages.

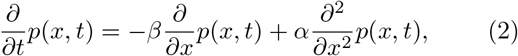

where the diffusion parameter is converted to *α* = *σ*^2^*/*2. The Fokker-Planck equation is a linear partial differential equation and conserves probability so that:

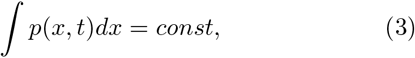

which allows us to solve it for any initial condition using Green’s function with the initial distribution in form of a Dirac delta function *δ*(*x*). The fundamental solution to the Fokker-Planck equation (Eqn. 2) is [45]:

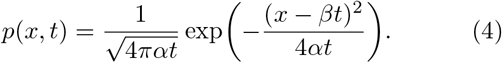

For non-zero drift and diffusion, this equation dictates that the average position of the population changes linearly with time and its standard deviation changes as the square root of time. Under our assumption that the distribution originates as a delta function prior to our observed data, we apply a shift of variable origins *x* →*x* −*x*_0_, *t* →*t*− *t*_0_ for some constant reference values of *x*_0_, *t*_0_.

Following the shift, we obtain the solution:

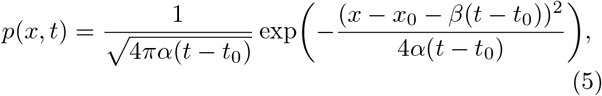

which describes a population experiencing constant drift and gradually widening diffusion or uncertainty. This probability distribution depends on four free parameters, which we group as *θ* = {*α, β, x*_0_, *t*_0_} and jointly infer from the observed data. We note that there are no physical restrictions such as non-negativity on the reference values {*x*_0_, *t*_0_}; since the drift-diffusion model (Eqn. 1) is only expected to apply in a certain age range with parameters {*α, β*} , the reference values parameterize the morphological distribution at the beginning of that age range.

### D. Maximum likelihood estimation

For model fitting, each patient in our dataset contributes one scan independently of the others by following a stochastic trajectory from birth to the age at scan time *t*_*i*_. The joint likelihood of the entire dataset is therefore given by the product of individual probabilities by independence:

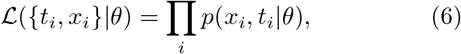

which we can now maximize to construct the maximum likelihood inference of the parameters:

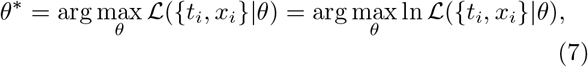

where the log-likelihood of the dataset of *N* data points is:

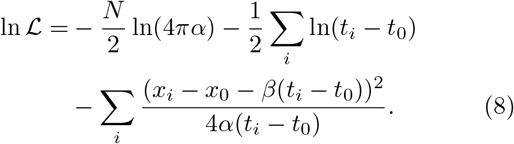

We fit the model by setting *∂* ln ℒ*/∂θ* = 0, deriving joint Fixed Point and Newton’s Method update equations, and iterating until convergence (Appendix F). To identify potential regime changes in the governing dynamics due to developmental and growth differences in children and adults, we conducted joint parameter estimation for sex-stratified cohorts that were further split with a floating age cutoff. Additionally, we fit models on sex-aggregated data to determine the impact of sex differences in the evolution of aortic size and shape. By partitioning our datasets across a range of age cutoffs, we compared the Bayesian information criteria (BIC) of the fitted models for each parameter: sex-stratified, age-split; sex-stratified, all ages; sex-aggregated, age-split; and sex-aggregated, all ages. Minimizing the BIC in this way allowed us to identify an optimal division of the dataset by age and sex and obtain fitted drift-diffusion models with the potential for age- and sex-driven dynamical regimes. If there was no age where fitting age-split models was advantageous over the all-age model, then a single drift-diffusion model was selected. If there was no advantage to divide the data by sex, the model was fit on sex-aggregated data (Appendix G, Fig. 5).

### E. Multivariate outlier detection

Given the functional form of Eqn. 5, the expected population mean trajectory is *x* = *x*_0_ + *β*(*t* − *t*_0_), and the standard deviation range is 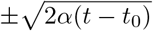 . This “envelope” of the expected stochastic population evolution defines a region of normal aortic morphology across the human lifespan. We expect that disease processes, such as AD, will result in morphology that is a statistical outlier, where observed measurements lie beyond the standard deviation range at a given age. In this section we extend the measurement of outliers to the multivariate case and turn it into a statistical threshold rule.

Following the fitting of separate models for each morphological parameter, we collect them into a multivariate model. The drift rate and reference parameters become vectors ***β*** = [*β*_1_, *β*_2_, · · ·]^⊤^; **x**_0_ = [*x*_0,1_, *x*_0,2_, …]^⊤^, the diffusion rate becomes a diagonal matrix **A** = *diag*([*α*_1_, *α*_2_, …]). The reference times are used to shift the time of scan *t*_*j*_ separately in each dimension 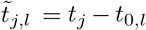, forming a vector 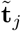. This collection of fitted parameters defines a multivariate Gaussian model in which the mean drifts uniformly and the covariance grows uniformly:

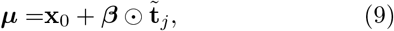

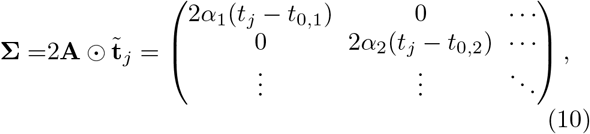

where the symbol ⊙ refers to elementwise product (Hadamard or Schur product), that reconciles the universal patient time stamp with morphological parameter– specific reference times.

Given the multivariate Gaussian model, we need to evaluate whether a scan with morphological parameters **x**_*j*_ constitutes a significant deviation from the mean. To do that, we compute the Mahalanobis distance [46]:

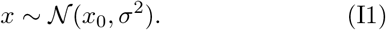

which is a multivariate extension of z-score, scaling distances in each direction through the covariance matrix. The *d*^2^ values of the multivariate reference distribution follow a chi-squared distribution with *m* degrees of freedom. To identify whether a given data point is an outlier with respect to the reference distribution for a 95% confidence level, we compare the data point’s associated *d*^2^ value to the complementary cumulative function of the chi-squared distribution evaluated at *χ*^2^(0.05, *m*). For our morphologic characterization of the aorta with *Ã* and 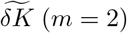, the threshold for outlier values of *d*^2^ is given by *χ*^2^(0.05, 2) ≈ 5.99, whereas for either of the univariate models it is *χ*^2^(0.05, 1) ≈ 3.84.

## III. RESULTS

### A. Stochastic modeling of normal aortic morphology

#### 1. Aortic size (Ã)

Model fitting of normalized thoracic aortic surface area was optimized by splitting the data at an age of 19 years for sex-stratified datasets (Fig. 5), characterizing two distinct regimes of morphologic evolution for each sex. During childhood (age *<*19 years), aortic surface area increases more quickly with drift rates of *β* = 0.0357, 0.0392 per year for females and males, respectively (Fig. 3, Table I). Following this expected period of somatic development, aortic surface area increases at a slower rate throughout the lifespan (*β* = 0.0064, 0.0110 per year for females, males). Notably, male aortas grow more quickly than female aortas at all ages, especially so for adults (≈70% faster), leading to an expected difference in normal aortic size for men and women that compounds over the lifespan.

**FIG. 3.**
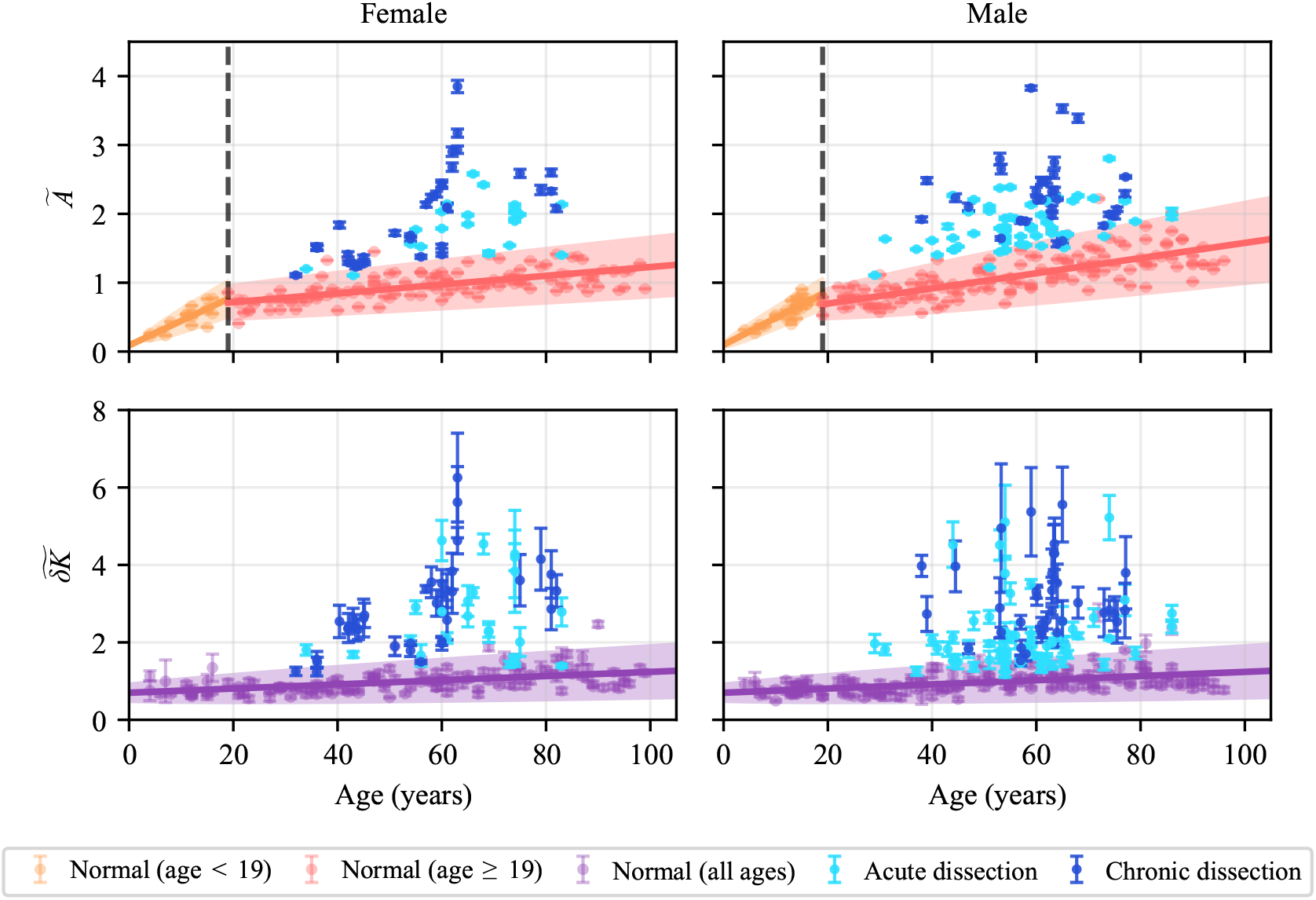
Stochastic drift-diffusion models of normalized thoracic aortic surface area (*Ã*) and fluctuation in integrated Gaussian curvature 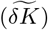 for patients with no aortic pathology. For surface area, child and adult models were fit separately for females and males, with an empirically derived cutoff at age 19. For 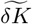, a single model was fit for patients of both sexes and all ages. See Appendix G and Figure 5 for optimization details. The mean expected growth trajectory for each model is plotted as a solid line, and the shaded region is the 95% confidence interval for normal aortic morphology. Sex-stratified data of normal patients underlie the population models. Held-out pathologic measurements of patients with acute and chronic dissections are shown in light and dark blue, illustrating the presence of aortic disease as a departure from the expected drift and diffusion of normal aortic morphology across the human lifespan.

**TABLE I.** Estimated initial conditions (*x*_0_, *t*_0_), drift (*β*) and diffusion (*α*) parameters for each fitted model.

| Model Parameters $\theta = \{\mathbf{x}_0, \mathbf{t}_0, \beta, \alpha\}$ [1, yr, yr<sup>-1</sup>, yr<sup>-1</sup>] | | | |
| --- | --- | --- | --- |
| | | $\tilde{\mathbf{A}}$ | $\tilde{\delta\mathbf{K}}$ |
| Female | Child | (0.1810, 2.5886, 0.0357, 0.0005) | (0.6346, -12.1986, 0.0054, 0.0005) |
|  | Adult | (0.4739, -18.1741, 0.0064, 0.0002) |  |
| Male | Child | (0.0811, -0.3896, 0.0392, 0.0003) |  |
|  | Adult | (0.5539, 7.0672, 0.0110, 0.0005) |  |

The diffusion parameter *α* quantifies the rate at which heteroscedastic variance accumulates (Table I) and is not directly comparable to the drift value because the distribution width grows as square root of the time in contrast to linear drift (Eqn. 5, Fig. 3). Similar to the trend observed for the drift coefficients, adult males also exhibit a greater amount of heteroscedastic variability than adult females, with a diffusion rate more than twice as large as their sex-stratified counterparts (Table I).

In addition to the normative measurements and fit models, pathologic measurements of normalized surface area for patients with acute and chronic thoracic ADs demonstrate the manifestation of aortic disease as a departure from the expected stochastic evolution of normal aortic size. Qualitatively, this departure appears as pathological points lying mostly outside the 95% confidence interval of normal patients; quantitative outlier detection is performed below.

#### 2. Aortic shape 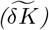

In contrast to aortic size, the optimization process for fitting the evolution of normalized fluctuation in integrated Gaussian curvature 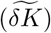 identified a single sex-aggregated, all-age drift-diffusion model as optimal for both sexes (Fig. 5). There was no age cutoff that resulted in better fit models, indicating that aortic shape belongs to a single dynamical regime, rather than evolving with distinct dynamical phases like aortic size (Fig. 3). Additionally, fitting a single model to all of the data was preferred to separate models for each sex, which demonstrates the sex- and therefore size-invariant nature of 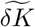.

Normalized fluctuation in integrated Gaussian curvature 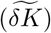 increases more slowly than surface area for all models, with drift rates of *β* = 0.0054 per year. The fitted diffusion rates are similar to those for aortic size, with *α* = 0.0005 per year (Table I). Together, this indicates the aorta approximately preserves its shape throughout a lifetime of normal growth, while patient heterogeneity increases with age, similarly to the aortic size.

The shape parameter 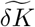 for patients with acute and chronic AD are also mostly outside of the 95% confidence interval of normative models, similarly to the surface area (Fig. 3); quantitative outlier detection for both parameters is performed below.

#### 3. Model validation

We validated the fitted models against held-out data using two complementary procedures. First, a 5-fold cross-validation procedure was performed for each sex, where drift-diffusion models were trained on 80% of the data and evaluated on the held-out 20% using the squared Mahalanobis distance to the multivariate 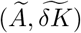 distribution. The hold-out distances match the expected *χ*^2^(2) distribution closely (Fig. 4); 95% of female and 97.2% of male hold-out points fall within the 95% confidence threshold (*d*^2^ = 5.99), indicating the predicted confidence interval neither underestimates nor overestimates the spread of unseen normal aortic data. Second, longitudinal fidelity was tested on followup scans from the *n* = 77 multi-scan patients whose later scans were held out during fitting; 93% of female and 96% of male follow-up scans fall within the 95% confidence threshold (Appendix J, Figs. 9, 10). A complementary analysis of follow-up trajectories in relative-time coordinates reveals that observed morphologic change at inter-scan intervals shorter than ≈1 month is dominated by CT-derived measurement noise rather than biological change (Fig. 11).

**FIG. 4.**
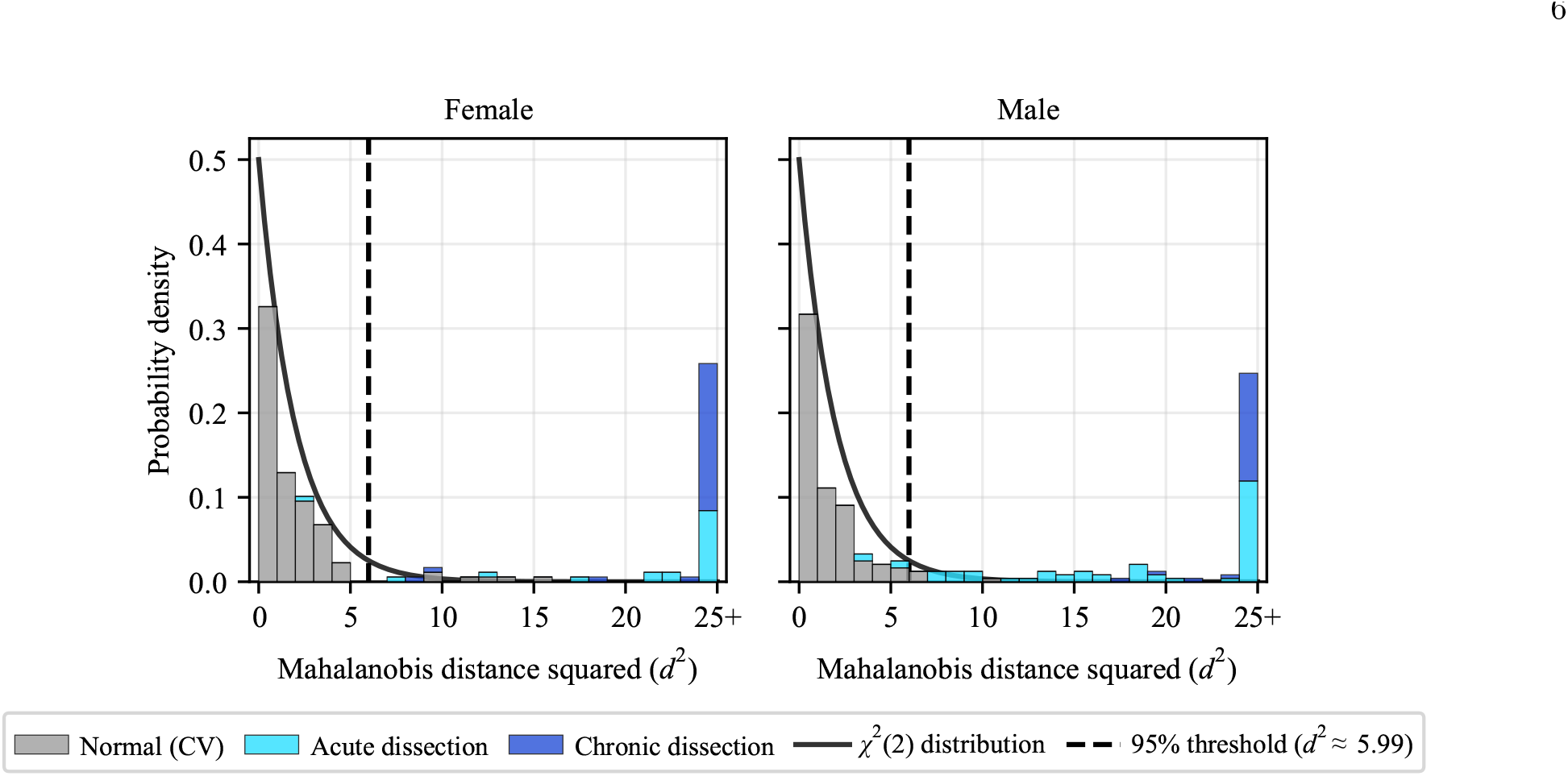
Sex-stratified distributions of *d*^2^ values calculated from morphologic measurements of 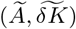 for: (1) all hold-out sets of a 5-fold cross validation of the normal cohorts, and (2) patients with acute and chronic thoracic ADs. For the normal cohorts, cross-validation yields empirical outlier rates of 5.0% (female) and 2.8% (male), consistent with the 5% rate expected under the null *χ*^2^(2) distribution (95% confidence threshold *d*^2^ ≈ 5.99). For the acute dissection cohorts, 95.7% of female and 93.7% of male data are identified as statistically significant outliers (*p <* 0.05). For the chronic dissection cohorts, 100% of the data are identified as statistical outliers for both sexes (*p <* 0.05). Together, these results validate the generalizability of the stochastic models to unseen normal aortic data, and they demonstrate the ability to accurately identify pathologic instances of aortic morphology with respect to the normative baseline of expected drift and diffusion across the human lifespan.

In addition to these validation tests for our drift-diffusion model, we also considered alternative models with fewer parameters. By comparing their fits by BIC, we determined the complexity of our 4-parameter drift-diffusion model is preferred to the other more parsimonious models (Appendix I).

### B. Discriminating dissection from normal morphology

Having validated models of normal aortic morphologic evolution, we next evaluated their ability to distinguish pathologic from non-pathologic aortic morphology. We applied the fitted models to additional datasets of patients with acute (*n* = 86) and chronic thoracic ADs (*n* = 70). For each diseased measurement of *Ã* and 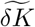, the Mahalanobis distance from the non-pathologic multivariate distribution was calculated and compared to the expected *χ*^2^ distribution (Fig. 4). As shown in Table II, 95.6% (100%) of female and 93.6% (100%) of male acute (chronic) dissections were identified as outliers (*p <* 0.05). When compared to the morphologic characterization of disease using *Ã* or 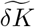 alone, the multivariate distribution exceeds the performance of either univariate model in both outlier detection and distributional separation from normal morphology (Tables II and III).

**TABLE II.** Outlier fractions identified by univariate and multivariate models in normal, acute dissection, and chronic dissection cohorts, stratified by sex. Bold values correspond to the best performing model for each dataset.

| Model | Female |  |  | Male |  |  |
| --- | --- | --- | --- | --- | --- | --- |
|  | Normal | Acute | Chronic | Normal | Acute | Chronic |
| Univariate: $\tilde{A}$ | 0.04 | 0.91 | **1.00** | 0.05 | 0.84 | 0.94 |
| Univariate: $\tilde{\delta K}$ | **0.02** | 0.83 | 0.91 | **0.04** | 0.79 | **1.00** |
| Multivariate: $(\tilde{A}, \tilde{\delta K})$ | 0.06 | **0.96** | **1.00** | **0.04** | **0.94** | **1.00** |

**TABLE III.** Median squared Mahalanobis distances from the non-pathologic distribution, computed for univariate and multivariate models in acute and chronic dissection cohorts, stratified by sex. The reference outlier threshold is *χ*^2^(0.05, 2) ≈ 5.99 for the multivariate model and *χ*^2^(0.05, 1) ≈ 3.84 for univariate models. Bold values correspond to the model with the greatest median squared Mahalanobis distance for each dataset.

| | | $\tilde{A}$ | $\tilde{\delta K}$ | $(\tilde{A}, \tilde{\delta K})$ |
| --- | --- | --- | --- | --- |
| **Female** | Acute | 21.4 | 18.2 | **53.3** |
|  | Chronic | 23.5 | 48.8 | **69.0** |
| **Male** | Acute | 9.56 | 11.6 | **19.4** |
|  | Chronic | 22.8 | 41.1 | **65.3** |

## IV. DISCUSSION

This clinical dataset presents an unusual setting for stochastic modeling. Many stochastic models are assumed to be Markovian so that the conditional distribution of an incremental step in *x* only depends on the current value of *x*. Consequently, inferring the parameters of such processes often relies on *local statistics* of increments [48]. In contrast, our cohort consists of a *collection of endpoints* of putative random walks observed at different times, so we infer the population-level drift and diffusion from the cross-sectional distribution of these endpoints. As such, this approach is more akin to stochastic generative modeling [52] than to local-statistics inference, and we are conceptually inspired by stochastic block models from network science [53, 54]. We leverage the existence of exact solutions to the Fokker-Planck equation [45], which are available for only a few forms of drift-diffusion dynamics but allow computational inference iterations to converge within seconds. More generally, the Fokker-Planck equation can be solved numerically within the inference loop, enabling more complex functional forms at the cost of computational resources.

### A. Normal aortic morphology is dynamic

Our models demonstrate that the normal aorta is not a static structure. Even in the absence of disease, aortic morphology changes continuously throughout life (Fig. 3, Table I). The mean of the population distribution shifts linearly in time at rate *β*, while its standard deviation grows as 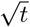, so even when *α* is small in absolute magnitude relative to *β*, its accumulated effect over the human lifespan is comparable to that of the drift. The growth trends show a strong sex difference in that adult male aortic growth in *Ã* outpaces that of adult females by approximately 70% (*β* = 0.0110 vs 0.0064 yr^−1^), and male inter-individual variability accumulates more than twice as fast (*α* = 0.0005 vs 0.0002 yr^−1^, Table I), producing a sex-based divergence in normal aortic size that compounds with age. Notably, across the adult lifespan of 20–100 years, the expected mean *Ã* shifts by about half of the 95% confidence interval width for females and a full interval width for males. Adult size and shape evolve at comparable rates, but the two metrics differ in their regime structure, as aortic size exhibits a childhood phase consistent with somatic development followed by slower adult growth, while aortic shape is governed by a single dynamical regime across all ages (Figs. 3, 5). Though prior normative studies have documented adult expansion in size [16–19] and the somatic-growth phase in children [14, 15] separately, the unified continuous-time framework here resolves both regimes in a single inference and reveals that no analogous transition occurs in shape.

**FIG. 5.**
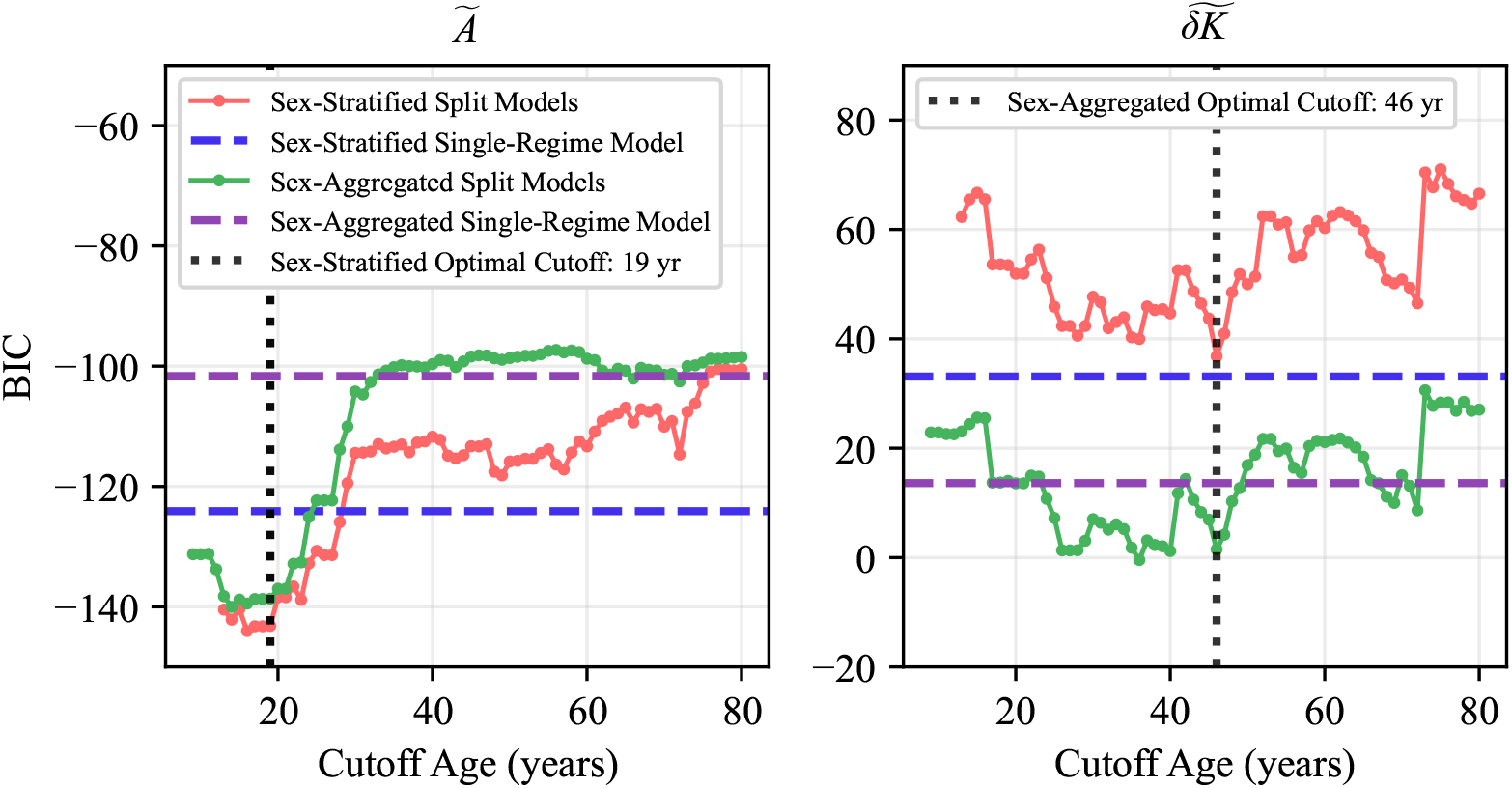
For *Ã*, the sex-stratified, age-split BIC is minimized at an age cutoff of 19 years. For 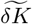, the sex-aggregated, age-split BIC is minimized at an age cutoff of 46 years. However, because this does not correspond to any known biological transition, instead of introducing a discontinuity in the mean trajectory we choose to select a sex-aggregated, single-regime model.

For both size and shape, the increasing heterogeneity captured by *α* admits a biological interpretation that individuals diverge from one another as risk factors for cardiovascular disease accumulate over time. Even when the population-mean trajectory is well-described by a linear deterministic drift, the accumulation of biological damage from established risk factors (including hypertension, dyslipidemia, and smoking history) is a plausible source of growing inter-individual variability [17, 55–58]. At the tissue level, our macroscopic findings are consistent with mechanisms of gradual morphological change reported in the aortic-wall literature; progressive elastin degradation, collagen remodeling, smooth muscle cell loss, and cumulative oxidative damage have all been described as being modulated by the local mechanical environment and systemic factors that vary across individuals [58–63]. While we do not directly validate any of these mechanisms in this study, the macroscopic dynamics we observe—slow drift in the population mean coupled with steady accumulation of inter-individual variability—are what one would expect at the population level from tissue-level processes proceeding at individual-specific rates. The sex-specific divergence in *β* and *α* for aortic surface area may partly reflect heterogeneous accumulation of the same cardiovascular risk factors over the lifespan rather than intrinsic biological sex differences. This interpretation is consistent with Beeche et al.’s observation of significant age-sex interactions in unscreened biobank cohorts but not in the subset they screened for cardiovascular risk factors [16–19] (Appendix C).

### B. Joint size-shape characterization improves disease detection

Across the dissection cohorts, the multivariate size-shape characterization consistently outperforms either univariate model on both the number of individual disease points identified as outliers (Table II) and the distance of the disease populations from the normal distribution (Table III). This demonstrates that *Ã* and 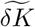 capture complementary aspects of the morphologic deviation introduced by aortic disease, which is mechanistically consistent with dissection introducing both size changes (false lumen expansion, focal aneurysmal degeneration) and geometric disruptions (flap formation, focal curvature deviation) [20, 55, 58]. While current clinical assessment reduces aortic morphology to a single scalar measurement of diameter, these results suggest that incorporating geometric descriptors of shape into the evaluation of thoracic aortic pathology provides a more complete characterization of the morphologic state of the aorta. However, not all shape metrics are equally informative; descriptors such as tortuosity index underperform 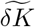 at distinguishing diseased morphology [20] (Appendix E).

The Mahalanobis-distance formulation generalizes from the two coordinates used here to arbitrarily many dimensions, in principle admitting additional features such as tortuosity [64] or hemodynamic descriptors like the blood pressure difference between the true and false lumens [65–68]. Introducing more metrics increases the Mahalanobis distance from the reference distribution, but it also raises the outlier threshold *χ*^2^(0.05, *m*), ensuring that the addition of more features improves detection only when the added features carry information not already captured by the existing coordinates [69]. The *χ*^2^(0.05, *m*) threshold enforces judicious feature selection, which is particularly important in clinical applications where many candidate features (imaging-derived metrics, lab values, comorbidities) compete for inclusion.

### C. A probabilistic envelope for clinically defining “normal”

Current clinical practice for identifying aortic disease defines “normal” anatomy using a static mean diameter of 30 mm [39, 70], a convention that reduces the richness of three-dimensional aortic morphology to a single scalar and that ignores age-related heterogeneity within the non-pathologic population. The stochastic framework presented here instead defines normal aortic size and shape as a probabilistic distribution that evolves with age and varies by sex, shifting the clinical perspective from a binarized view of normal to a probabilistic one. The expected mean and the uncertainty around it are both non-stationary, so thresholds derived from a static mean become increasingly misaligned with the true normal baseline as patients age.

Three clinical implications follow. First, for diagnostic evaluation, age- and sex-specific distributions allow individual scans to be flagged on exit from a given confidence interval of normal morphology, supporting earlier identification of pathologic deviation than is possible with fixed, binarized thresholds. Second, for pre-operative decision-making, the framework provides a principled basis for revisiting current surgical thresholds. Sex-specific diameter thresholds are already accepted for abdominal aortic aneurysms (50 mm for women, 55 mm for men [39]), but the descending thoracic aorta retains a single 60 mm threshold that derives from twice an assumed “normal mean” of approximately 30 mm [39, 41, 70]. The current threshold for descending thoracic aneurysm and type B dissection repair derives largely from observational studies of adverse aortic events [39, 41, 42, 71], rather than from a calibrated model of when the aorta has departed from normal—a gap that the probabilistic envelope established here directly addresses by replacing the assumed static normal with a fitted, age- and sex-stratified one. Third, for post-operative monitoring, this work reframes what “returning to normal” means. A common treatment of thoracic aortic dissection is thoracic endovascular aortic repair (TEVAR), in which a stent is inserted into the aorta to confine blood flow within itself and depressurize the wall, allowing the aorta to remodel toward the normal shape [27, 30, 31]. Because the probabilistic envelope of normal itself evolves with age, what constitutes successful post-operative remodeling is not a fixed target but a dynamic one.

Since individual aortas may deviate from the population envelope in patient-specific ways, understanding post-operative morphologic change requires both a population-level target distribution and models of individual trajectories. The Fokker-Planck framework presented here infers the former from single-timepoint cross-sectional scans, which is the regime where most clinical data are available. Sparse identification of nonlinear dynamics (SINDy) [4, 49], applied in [47] to post-operative abdominal aortic aneurysm remodeling, infers patient-specific Ordinary Differential Equations (ODEs) from longitudinal sequences of patient imaging. This combination is general, as any morphological system observable through cross-sectional or longitudinal imaging admits both descriptions, with the choice of method determined by the available data rather than the system being studied. Integrating the two into a unified outcome-specific framework, in which patient-specific dynamical trajectories are evaluated against the age- and sex-stratified normative distributions established here, is a natural direction for future work.

### D. Limitations and future directions

Our analysis relies on simplifying geometric, dynamical, and clinical assumptions that inform future studies. *Cohort:* The demographics of our normal population were skewed, with 78.8% of female and 64.9% of male patients identifying as Black, reflecting the trauma population admitted to the University of Chicago and the predominantly Black population on the south side of Chicago. Body surface area, a common normalizing variable for aortic dimensions, was not available because reliable height and weight measurements are frequently absent in the trauma setting. The cohort is single-center, with limited longitudinal coverage owing to the typical course of care for trauma patients, and is not screened for cardiovascular risk factors, so the sex-specific divergence in *β* and *α* captured here cannot be fully separated from accumulated risk-factor effects. Future datasets with more balanced demographics, richer longitudinal sampling, and risk-stratification would clarify the generality of the findings reported here. *Pathology scope:* Only acute and chronic dissections are tested in this study. Other pathologies such as thoracic aortic aneurysms and abdominal aortic disease are natural candidates for further analysis under the same framework, as are non-aortic morphological systems where serial or cross-sectional imaging captures organ-scale remodeling. *Dynamics:* Modeling aortic morphologic evolution as a constant drift-diffusion system is a simplifying assumption that provides interpretability and parametric parsimony. Incorporating patient-specific covariates, nonlinear dynamics, multivariate evolution of correlated morphological parameters, pre-operative natural course of aortic disease, or time-varying parameters could refine the framework or extend its applicability to other anatomic systems.

## V. CONCLUSION

This study establishes a continuous-time stochastic framework for normal thoracic aortic morphology across the human lifespan, with inter-individual variability treated as a fitted diffusion parameter *α* rather than as residual scatter. The fitted sex-stratified for normalized aortic size envelopes capture two distinct regimes— childhood expansion followed by adult growth, with the developmental-to-adult transition near age 19 identified directly by BIC—and the fitted sex-aggregate identifies a single regime for shape shared by both sexes, with heteroscedastic variability accumulating at a rate comparable to the population-mean drift over the lifespan. Applied to independent cohorts of thoracic aortic dissections, the same envelope that describes normal aging identifies over 95% as statistical outliers, with disease detection thus serving as an out-of-distribution test of the framework. This approach mathematically supports a shift in aortic-pathology assessment toward dynamic, age- and sex-stratified probabilistic baselines, and combines naturally with longitudinal methods that infer individual-trajectory dynamics, opening a route to physically interpretable models of organ-scale remodeling wherever serial or cross-sectional imaging is available.

## Data Availability

All data produced are available online at https://github.com/SurgBioMech/stochastic-normals

## ACKNOWLEDGMENTS

This work was supported by the National Institutes of Health, USA, NHLBI Grant R01-HL159205 (to LP) and Medical Scientist Training Program Grant T32-GM007281 (to MM). The Center for Research Informatics is funded by the Biological Science Division, USA at the University of Chicago.

Author contributions are as follows. **Conceptualization:** LP, AK, MM, KT, JP. **Data Curation:** KT, KY, MM, JP, LP. **Formal Analysis:** MM, JP. **Funding Acquisition:** LP. **Investigation:** MM, KT. **Methodology:** AK, LP. **Project Administration:** LP. **Resources:** LP. **Supervision:** LP, AK. **Validation:** MM. **Writing – Original Draft:** MM, JP, KT. **Writing – Review & Editing:** LP, AK, MM, JP.

## Appendix A: Code availability

The data and code used for pre-processing, model construction, and analysis are publicly available at https://github.com/SurgBioMech/stochastic-normals.

## Appendix B: Study cohorts

Patients without aortic pathology were identified through retrospective chart review of individuals who presented to the University of Chicago Trauma Center and the Comer Children’s Hospital Trauma Center between January 1, 2010 and December 31, 2023. Because trauma patients routinely undergo CTA of the chest, this population provided a practical source of aortic imaging in individuals without known aortic disease. Patients with any form of aortic pathology or a history of connective tissue disorder were excluded, and patients were selected to balance sex and age groups.

CTA scans from patients with aortic pathology were obtained from the University of Chicago and from deidentified imaging datasets provided under data use agreements between the University of Chicago and Medtronic plc (Mounds View, Minnesota), and between the University of Chicago and W. L. Gore & Associates (Newark, DE). For acute dissection data, 40 scans from 33 people (10 female, 23 male) were obtained from the GORE TAG 08-01 dataset [72], 41 from 38 people (7 female, 31 male) were obtained from the Medtronic dataset, and 18 scans from 13 people (5 female, 8 male) were obtained from the University of Chicago CARES registry. For the pre-operative chronic dissection data, 57 scans from 23 people (10 female, 13 male) were also obtained from the University of Chicago CARES registry. Sourcing the dissection cohort across three independent clinical sites and imaging protocols provides a stronger test of generalization than a single-institution cohort would, at the cost of inter-protocol acquisition variability.

All data collection and analyses were conducted in accordance with the Declaration of Helsinki and approved by the University of Chicago Institutional Review Board (IRB20-0653, IRB21-0299).

## Appendix C: Comparison to prior normative cohorts and studies

Prior normative studies of aortic morphology consistently report slow age-related dilation and sex differences in adult aortic size [16–19], but they share two structural limitations. First, inter-individual variability is treated as a residual. Beeche et al. report descriptive statistics within 10-year age strata and use sex-adjusted linear regression models [16]; Redheuil et al. use 10-year bins and ANOVA F-tests for trend across bins [17]; Rylski et al. use continuous linear regression and compare young (*<*45) and senior (*>*70) groups categorically [18]; and the DANCAVAS-derived thresholds discussed by Elefteriades and Beckman are defined cross-sectionally within a single age bin spanning 15 years [19]. These approaches share the assumption that the population mean evolves linearly or piecewise-linearly with age, with interindividual variability reported as a residual standard deviation rather than as a dynamical quantity. Our framework instead treats *α* as a fitted parameter governing how the population distribution widens over time, and shows that this heteroscedasticity is comparable in magnitude to the mean drift itself over the human lifespan. Second, prior cohorts lack the lifespan coverage to resolve developmental-to-adult regime structure. Rylski’s cohort had no pediatric subjects; Redheuil’s youngest stratum began at age 20; the UK Biobank normative subgroup is bounded below at 37; and DANCAVAS only covered the ages of 60-74. The continuous-time formulation our framework adopts uses every patient scan to inform a parsimonious parameter set, avoids the discontinuities that age-bin boundaries introduce, and produces lifespan envelopes from a single inference procedure rather than a stack of independently fit bin-specific cutoffs.

Our cohort of 311 patients (388 scans) is substantially larger than the Redheuil and Rylski cohorts and orders of magnitude smaller than the biobank-scale studies, but differs in three coordinated features that together enable the analyses presented here. Lifespan coverage from 4–99 years with approximately balanced representation across each decade (Table IV) is what makes the BIC-driven identification of a developmental-to-adult regime change near age 19 possible (Figs. 3, 5). Sex balance is enforced within each age stratum instead of only in aggregate, which makes the sex-specific drift and diffusion parameters in Table I credible because aggregate balance is consistent with strong age-by-sex confounding (the UK Biobank normative subgroup is approximately 71% female overall, and although Rylski’s cohort is near 50/50 in aggregate, age is modeled continuously without per-stratum sex balance being reported). Finally, sourcing patient data from a trauma center decouples the imaging indication from the structure being studied, because chest CTA in trauma is ordered by mechanism of injury rather than by suspicion of aortic or cardiovascular disease. This contrasts with Rylski’s cohort, which is dominated by chest-pain evaluation (84%) alongside smaller fractions of trauma and cancer staging, with the clinically-driven Penn Medicine Biobank, and with the UK Biobank normative subgroup, which is selected on metabolic and lifestyle criteria that correlate with vascular structure. Trauma sourcing carries its own biases, including the demographic skew of the population served by the University of Chicago (Section IV D), but the decoupling between imaging indication and aortic anatomy is a structural feature that prior cohorts do not share. A consequence of trauma sourcing is that our cohort, unlike Beeche’s normative UK Biobank subgroup, is not screened for hypertension, smoking, dyslipidemia, or glycemic dysregulation. In this way, it is closer to their overall UK Biobank and Penn Medicine Biobank cohorts. Beeche et al. found significant age-sex interactions in the overall cohorts but none in the normative subset, which suggests that the sex-specific divergence in adult drift and diffusion captured by *β* and *α* for aortic surface area may partly reflect the accumulation of cardiovascular risk over the lifespan rather than intrinsic biological sex differences. This is consistent with our interpretation of *α* as a population-level signature of heterogeneous risk-factor accumulation (Section IV A).

**TABLE IV.** Stratification of normal patients by sex and age group.

| Age Group (Years) | Unique Patients |  |
| --- | --- | --- |
|  | Female | Male |
| 0–9 | 5 | 6 |
| 10–19 | 14 | 28 |
| 20–29 | 15 | 17 |
| 30–39 | 15 | 20 |
| 40–49 | 16 | 18 |
| 50–59 | 15 | 21 |
| 60–69 | 16 | 18 |
| 70–79 | 15 | 18 |
| 80–89 | 16 | 18 |
| 90–99 | 10 | 10 |
| **Total** | **137** | **174** |

These design choices enable the central findings of our stochastic framework. Adult males grow approximately 70% faster than adult females in *Ã* and accumulate interindividual variability more than twice as quickly (Table I); the developmental-to-adult transition at age 19 is identified directly rather than imposed; and the heteroscedastic structure of normal aortic aging emerges as a fitted physical quantity (*α*) rather than being reported as residual scatter. None of these results are accessible from a cohort that lacks lifespan coverage, per-stratum sex balance, or sufficient parametric flexibility to model variance as a function of age.

## Appendix D: Size metric selection

The choice of the scalar descriptor used to represent aortic size is non-trivial, and each of the commonly available options—maximum diameter (*D*_max_), mean diameter 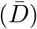, surface area (*A*), and volume (*V*)—carries biases that propagate into any downstream modeling. Maximum diameter remains the clinical standard and underpins current surgical thresholds [39, 41] because it is directly measurable from an axial CTA slice without further postprocessing, but this operational simplicity masks a well-documented sensitivity to the choice of measurement plane and to inter-observer variability [25, 73]. Mean diameter, obtained by averaging cross-sections along a reconstructed centerline, mitigates the plane-alignment bias but remains a one-dimensional reduction of a three-dimensional object. Surface area and volume, in contrast, are genuinely three-dimensional metrics: they are insensitive to axial plane orientation, naturally accommodate non-circular cross-sections, and integrate the contributions of tortuosity and length. Their tradeoff is that *A* and *V* inherit the variability of the upstream segmentation pipeline, most consequentially the choice of system boundaries at the iliacs, renals, celiac trunk, or supra-aortic branches, where small landmark shifts translate directly into large shifts in *A* or *V* and to a much lesser extent in *D*_max_ or 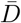 .

In this analysis, the tightly controlled boundary conventions described in Section II B suppress this source of noise such that *Ã* can be used confidently as the size coordinate in the drift-diffusion framework. We emphasize that this reliability is a property of the curated cohort and pre-processing pipeline rather than of the metric itself, and translating the framework to less controlled settings would require either comparable boundary standardization or explicit modeling of boundary-induced variance. The choice between *D*_max_, 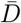 , *A*, or *V* is also less consequential for the inferred dynamics, as our previously published analysis of the thoracic aorta [20] demonstrated, these four descriptors collapsed onto one another with near-linear scaling, so the drift and diffusion parameters reported in Table I are expected to transfer to diameter-based analyses after appropriate rescaling. However, equivalence under linear rescaling is not equivalence in information content: *A* and *V* are integrals over the full segmented manifold, while *D*_max_ and 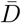 reduce it to a single plane or a one-parameter family. We therefore argue that when segmentation boundaries can be standardized, surface area or volume should be adopted as the preferred scalar descriptor for downstream morphologic modeling.

## Appendix E: Comparison of 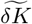 and tortuosity index as shape descriptors of aortic disease

Tortuosity index (TI), typically defined as the ratio of the centerline arc length to the chord length between two anatomical landmarks, is a widely used scalar descriptor of vessel geometry in studies of aortic disease [20, 74–76]. Khabaz et al. previously demonstrated that 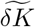 outperforms TI as a predictive metric of post-intervention outcome in aortic dissection [20]. Here we provide complementary statistical evidence that 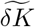 better captures the morphologic signature of aortic disease across the lifespan using the Mahalanobis-distance outlier framework used in the main text. We note that not all data in the main text have tortuosity indices measured, so this analysis relies on the subset that have all three morphologic descriptors available (*n* = 264 normal aortas; *n* = 28, 53 acute and chronic dissections).

Table V reports the fraction of data identified as statistically significant outliers (*p <* 0.05) under five model configurations: *Ã* alone, 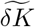 alone, 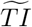 alone; the bivariate models (*Ã*, 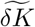), (*Ã*, 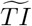) and 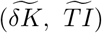; and the trivariate model 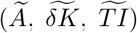. As a stand-alone shape metric, 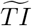 fails almost entirely, as it identifies 0% of female acute dissection cases and only 5% of male acute cases as outliers, with chronic detection rates of 9% (female) and 25% (male). In contrast, 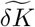 alone identifies 86% and 76% of female and male acute dissections and 90% and 97% of female and male chronic dissections, respectively.

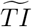 also fails as a replacement shape metric when combined with size. The 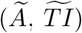 model identifies only 43% of female and 81% of male acute dissections as outliers, versus 86% and 100% for 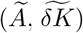. The chronic dissection detection rates exhibit the same pattern, with 67% and 94% for 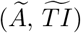 versus 100% for 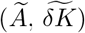 in both sexes.

**TABLE V.** Outlier fractions identified by univariate and multivariate models in normal, acute dissection, and chronic dissection cohorts, stratified by sex. Combinations including normalized tortuosity index 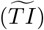 exhibit performance degradation for the identification of disease compared to using 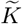 as a shape metric. Additionally, including 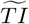 as a second shape metric in addition to *Ã* and 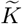 offers no improvement in disease detection, as well as loss of performance in female chronic dissection detection, when compared to *Ã* and 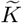. Bold values correspond to the best performing model for each dataset.

| Model | Female |  |  | Male |  |  |
| --- | --- | --- | --- | --- | --- | --- |
|  | Normal | Acute | Chronic | Normal | Acute | Chronic |
| $\tilde{A}$ | 0.04 | 0.43 | 0.76 | 0.06 | 0.95 | **1.00** |
| $\delta\tilde{K}$ | **0.03** | **0.86** | 0.90 | **0.04** | 0.76 | 0.97 |
| $\tilde{TI}$ | **0.03** | 0.00 | 0.09 | **0.04** | 0.05 | 0.25 |
| $(\tilde{A}, \delta\tilde{K})$ | 0.05 | **0.86** | **1.00** | 0.06 | **1.00** | **1.00** |
| $(\tilde{A}, \tilde{TI})$ | 0.05 | 0.43 | 0.67 | 0.06 | 0.81 | 0.94 |
| $(\delta\tilde{K}, \tilde{TI})$ | 0.05 | **0.86** | 0.91 | **0.04** | 0.67 | **1.00** |
| $(\tilde{A}, \delta\tilde{K}, \tilde{TI})$ | 0.04 | **0.86** | 0.95 | 0.06 | **1.00** | **1.00** |

Finally, the trivariate model 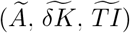 offers no improvement (as well as performance degradation for female chronic dissections) compared to the bivariate 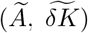 across all dissection cohorts. Adding 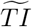 as a third feature does not improve performance because the additional discriminative information 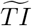 contributes is insufficient to offset the stricter trivariate threshold *χ*^2^(0.05, 3) ≈ 7.81. This illustrates of the principle of parsimony in feature selection [43, 44], as an uninformative feature does not simply fail to help, but it actively worsens detection by inflating the model’s null-distribution threshold without providing a compensatory increase in the signal carried by diseased cases.

Table VI reports the median squared Mahalanobis distance from the normal distribution for each model configuration. The pattern mirrors the outlier-detection results. As a stand-alone metric, 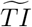 yields median distances below the univariate threshold of *χ*^2^(0.05, 1) ≈3.84 for all cohorts. In contrast, 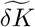 produces median distances well above the threshold in every cohort (9.67— 34.3). Substituting 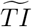 for 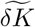 in the bivariate 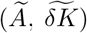 model again worsens the separation. Adding 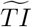 to 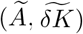 as a third feature only minimally increases every cohort’s median squared Mahalanobis distance, as there is little discriminative power that the additional shape metric offers.

**TABLE VI.** Median squared Mahalanobis distances from the non-pathologic distribution, computed for univariate and multi-variate models in acute and chronic dissection cohorts, stratified by sex. The reference outlier thresholds are *χ*^2^(0.05, 1) *≈* 3.84 for the univariate models, *χ*^2^(0.05, 2) *≈* 5.99 for the bivariate models, and *χ*^2^(0.05, 3) *≈* 7.81 for the trivariate model. Similar to the results of Table V, combinations including normalized tortuosity index 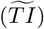 exhibit smaller distances for the identification of disease compared to using 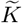 as a shape metric. Inclusion of 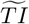 as an additional shape metric with *Ã* and 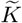 minimally increases distance compared to *Ã* and 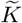 alone for disease cohorts, reflecting the low discriminative power of 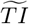.

| | | $\tilde{A}$ | $\delta\tilde{K}$ | $\tilde{TI}$ | $(\tilde{A}, \delta\tilde{K})$ | $(\tilde{A}, \tilde{TI})$ | $(\delta\tilde{K}, \tilde{TI})$ | $(\tilde{A}, \delta\tilde{K}, \tilde{TI})$ |
| --- | --- | --- | --- | --- | --- | --- | --- | --- |
| **Female** | Acute | 2.08 | 16.9 | 0.90 | 18.6 | 2.24 | 17.3 | 18.6 |
|  | Chronic | 9.75 | 34.3 | 0.57 | 42.6 | 12.5 | 34.5 | 45.7 |
| **Male** | Acute | 8.98 | 9.67 | 0.28 | 16.1 | 9.24 | 9.96 | 16.5 |
|  | Chronic | 27.2 | 31.5 | 1.38 | 58.0 | 29.3 | 33.3 | 62.3 |

Together, these results establish that 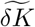 is not interchangeable with tortuosity index as a scalar shape descriptor of aortic morphology and is superior in discriminating pathologic aortic shape from the expected normal variation. 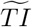 underperforms 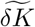 both alone and in combination with *Ã*, and including 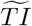 alongside 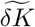 increases distance less than the associated increase of the 95% threshold. Further, this statistical method can be extended to accept or reject additional proposed clinical and morphologic features in future work.

## Appendix F: Maximum likelihood fitting for stochastic models

Since the likelihood is a differentiable function of four continuous parameters, we maximize it by setting *∂* ln ℒ*/∂θ* = 0 and solving for each parameter in terms of others. The resulting equations are coupled, so they can be solved by joint fixed point iteration where *q* is the iteration counter:

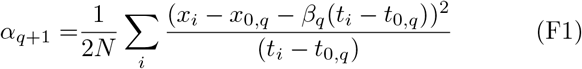

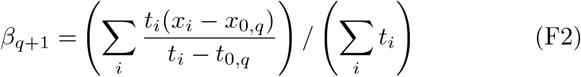

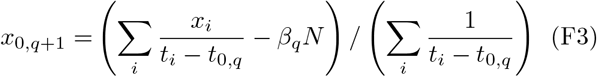

In contrast, the remaining equation for *t*_0_ does not allow an easy closed-form solution that would exclude *t*_0_ from the right hand side. We instead iterate it with Newton’s method:

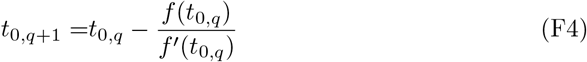

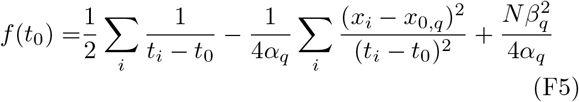

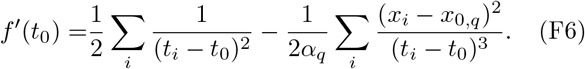

These inference equations depend on a variety of statistical summaries of the data, such as sample means, variances, covariances, and some higher moments. To facilitate convergence, we choose the following initial guess by assuming that the growth rate is not very high and reference age is close to 0 years:

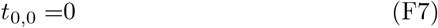

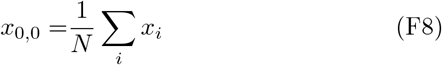

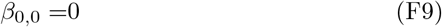

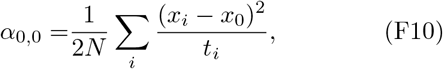

which is simple to compute directly from summary statistics but is not a fixed point of the iterations.

## Appendix G: Optimization of drift-diffusion models

To fit drift-diffusion models on our morphologic data, we implemented a joint optimization process for both male and female data simultaneously. For increments of 1 year, we applied a cutoff to divide the data into two subsets for each sex. A separate drift-diffusion model was fit on each subset of data, and the combined BIC of the four resulting models was computed at every candidate age cutoff (Eqn. G1).

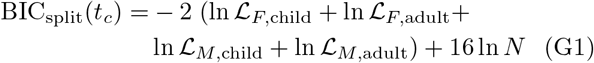

This was compared to the combined BIC of a single model for each sex that was fit on the entire respective datasets (Eqn. G2).

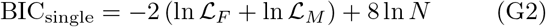

If the combined BIC_split_ for the candidate age-cutoff models was not lower than the combined BIC_single_ for the models fit on the full datasets, a single model was fit on each sex’s data. Otherwise, we identified the age cutoff that minimized the BIC_split_ and adopted a cut-off of the *latest* integer age within ΔBIC_split_ = 3 of this minimum. Differences in BIC of ≤3 are conventionally considered weak evidence against the more complex model [77], so all such candidate ages in this range are approximately equivalent in their statistical support among them. Therefore, we adopt the latest to maximize the number of patients assigned to the child model and improve the statistical power of the child-parameter inference.

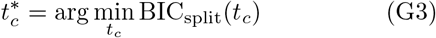

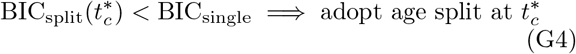

For *Ã*, two models for each sex with an age cutoff of 19 years minimized BIC_split_, while for 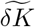, a single all-ages model for each sex was preferred (Fig. 5).

Additionally, we fit models on sex-aggregated data to determine if sex-stratification was optimal. As shown in Figure 5, sex-stratification and age-split models are preferred for surface area. However, for 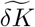, BIC identifies a sex-aggregated and age-split model at age 46 as optimal. Because this cutoff does not correspond to any known biological transition and would introduce a discontinuity in the predicted population mean, we instead adopt the sex-aggregated, single-regime model for both sexes.

## Appendix H: Stochastic model selection

Two alternatives to the 4-parameter drift-diffusion model—binned strategies with mean and variance estimated separately within age decades, and reduced functional forms with fewer free parameters—warrant explicit consideration. The first is rejected for the same reasons we identified in prior bin-based studies (Appendix C): bin-boundary discontinuities, statistical power lost to per-bin fitting, and the parsimony penalty of fitting dozens of independent parameters in place of four. The second alternative consists of a 2-parameter model describing the population by a fixed mean and variance (“MeanVar”), and a 3-parameter model adding a linear drift while still assuming a time-independent variance (“DriftVar”, Appendix I). Both admit closed-form maximum likelihood solutions and were evaluated under the same joint cutoff-optimization framework as the 4-parameter model.

To compare the performance of the 2- and 3-parameter models with our 4-parameter drift-diffusion model, we implemented sex-stratified and sex-aggregated optimization methods of BIC for each model (adjusted for the change in parameters), similar to the method described in Appendix G. Fig. 6 shows the optimization results, while Figs. 7 and 8 display the optimized 2- and 3-parameter models overlaid on the full dataset. For surface area, the sex-stratified drift-diffusion model with an age-cutoff of 19 years yields comparable fit to the sex-stratified 3-parameter “DriftVar” model at a similar cutoff. However, the optimized cutoff identified for the “DriftVar” model (age 38), does not correspond with the known developmental transition between childhood and adulthood. Therefore, we posit the drift-diffusion model is superior, as it implicitly models this biological transition while providing the ability to capture increasing heterogeneity in the aging population.

**FIG. 6.**
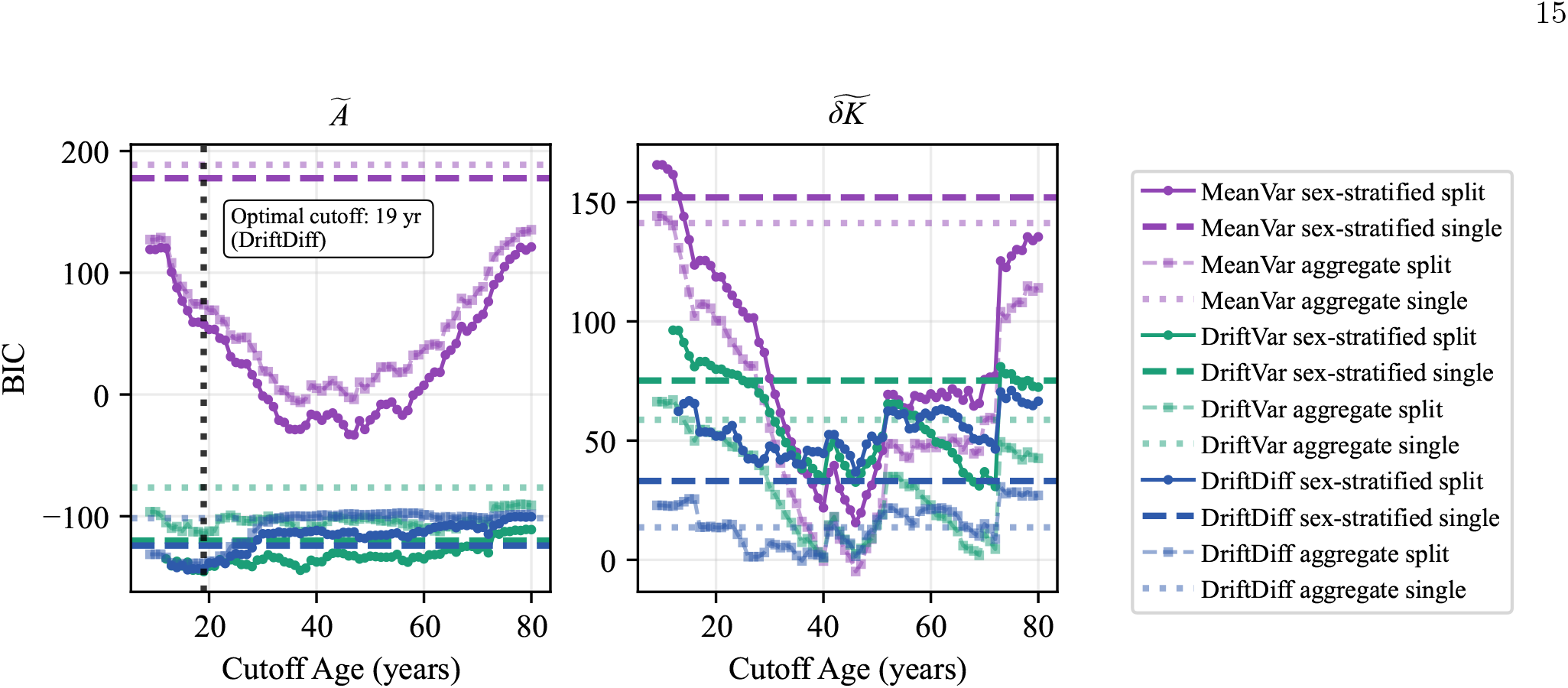
BIC calculated for 2-, 3-, and 4-parameter models of both male and female age-split data, as well as all-age data, for normalized thoracic aortic surface area (left) and normalized fluctuation in thoracic aortic curvature (right).

**FIG. 7.**
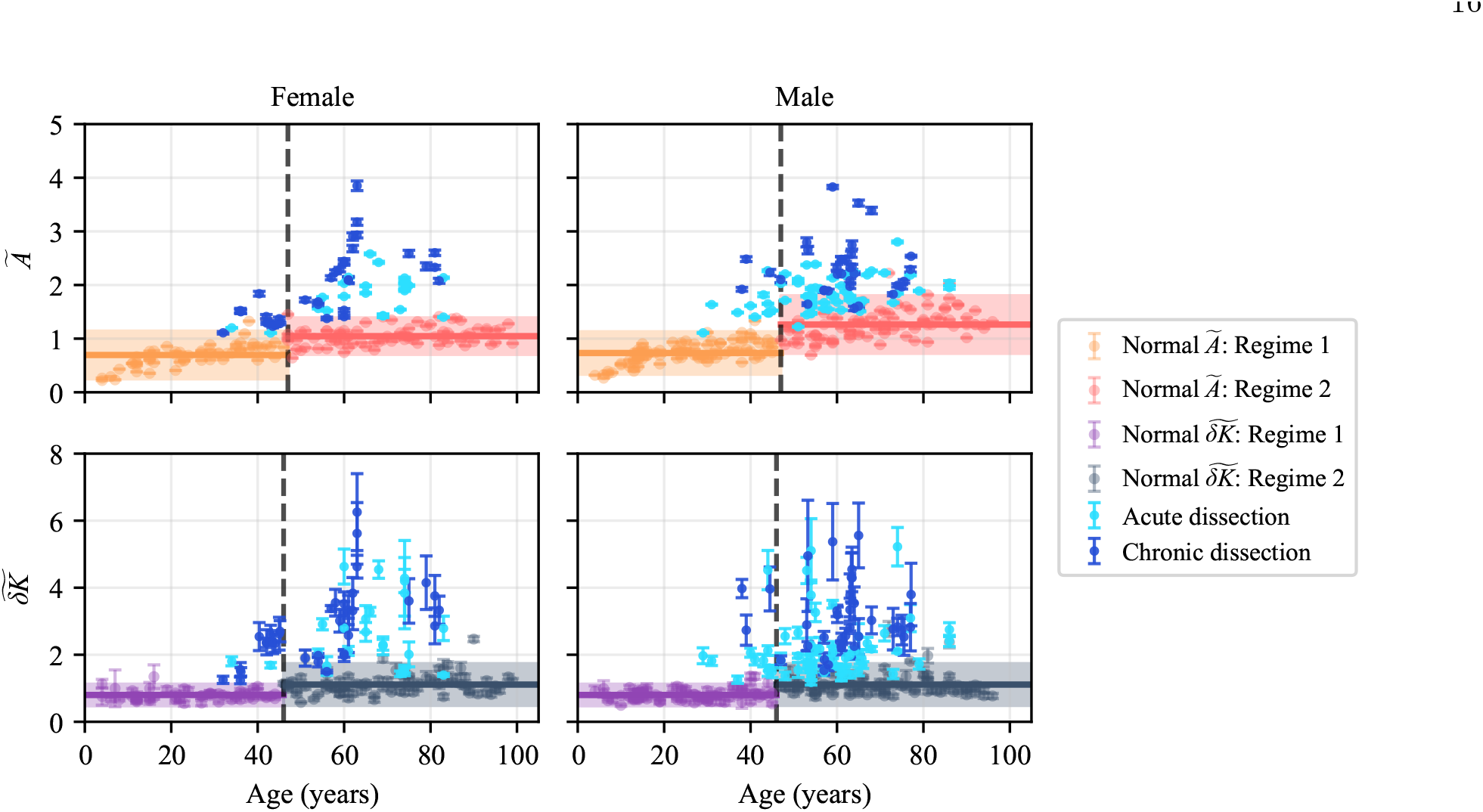
BIC-optimized 2-parameter MeanVar models of *Ã* and 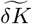.

**FIG. 8.**
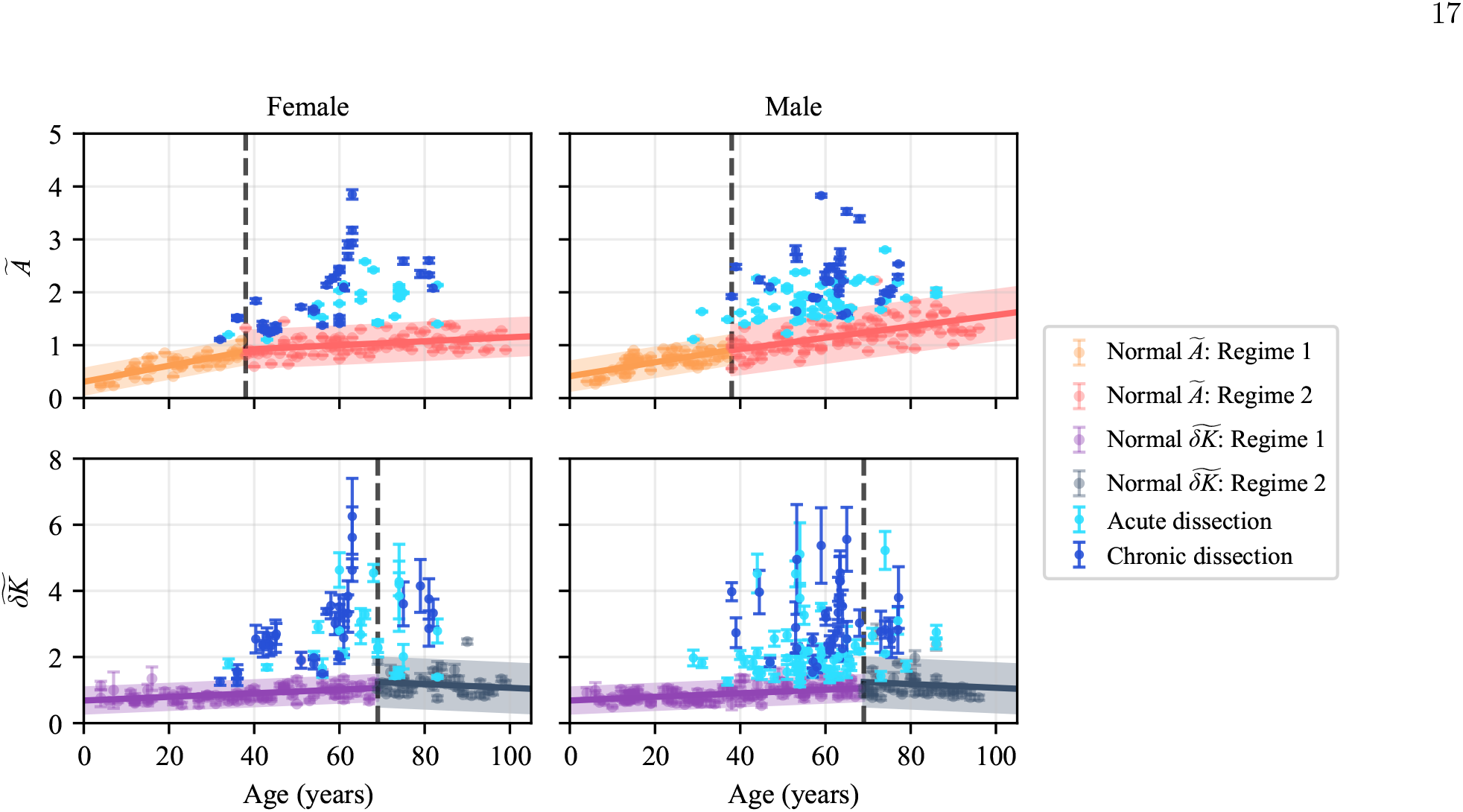
BIC-optimized 3-parameter DriftVar models of *Ã* and 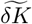.

For 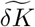, BIC marginally favors a sex-aggregated 2-parameter model with a mid-40s age cutoff, but this does not correspond to any known biological transition and would introduce a discontinuity in the predicted population mean. Similarly, the 3-parameter model is optimized by a sex-aggregated fit with an age cutoff of approximately 70 years, again introducing discontinuity and not corresponding with a biological transition. Both sex-aggregated drift-diffusion models outperform the sex-stratified drift-diffusion models at every age. While fitting with an age cutoff strictly improves BIC, we adopt the single-regime model and accept a small BIC penalty in exchange for the biological interpretability of 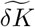 as a near-invariant shape metric across the lifespan. With these criteria, we are confident in our decision to select a single-regime, sex-aggregated 4-parameter drift-diffusion model for 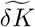 (Fig. 3).

## Appendix I: Derivation of simpler stochastic models

While we chose to fit a 4-parameter drift-diffusion model to our data, reduced Gaussian models with 2 or 3 parameters were also investigated as more parsimonious options. The two models both assume homoscedastic variance in contrast to the full drift-diffusion model whose variance grows with the square root of time. Because of this simplifying assumption, both models have closed-form MLE solutions that do not require iterative techniques to solve.

First, we derive the solutions for a 2-parameter model with no time dependence (“MeanVar”), where every observation of *x* is assumed to be drawn from a fixed Gaussian with unknown mean *x*_0_ and unknown variance *σ*^2^:

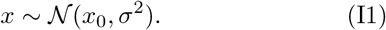

The MLE is obtained by setting *∂* ln ℒ*/∂θ* = 0 and solving for each parameter. For *θ* = {*x*_0_, *σ*^2^}, we obtain:

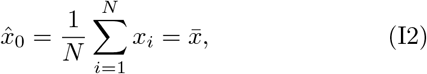

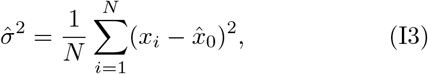

and find that the MLE for *x*_0_ is the sample mean and *σ*^2^ is the uncorrected sample variance, computed using the MLE of *x*_0_. In other words, the 2 parameter model provides a single, static estimate of the population’s mean and variance regardless of age.

Next, we derive the solutions for a 3-parameter model (“DriftVar”) that incorporates a linear drift term with the mean while keeping variance constant in time:

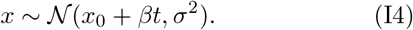

The parameter set is now *θ* = {*x*_0_, *β, σ*^2^}, and once again we set *∂* ln ℒ*/∂θ* = 0 and solve for each variable, which gives us:

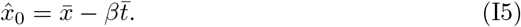

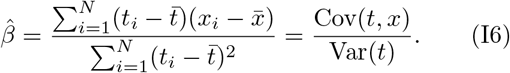

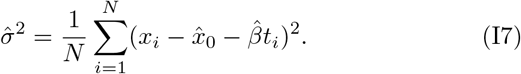

The MLE estimators for *x*_0_ and *β* in the 3-parameter model are identical to those obtained from ordinary least squares regression of *x* on *t*. This equivalence is due to the assumption of constant Gaussian noise, so that the log-likelihood reduces to a constant minus the sum of squared residuals (up to a *σ*^2^ scaling). Therefore, maximizing the likelihood and minimizing the squared error become the same optimization problem. The MLE also provides an estimate of *σ*^2^, in contrast to ordinary least squares which treats *σ*^2^ separately as a residual variance.

## Appendix J: Model validation

### 1. Cross-sectional validation

To validate our stochastic models of normal aortic size and shape, we conducted a 5-fold cross validation procedure using a two-dimensional Mahalanobis distance from the multivariate 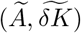 distribution. This two-dimensional space allows us to combine aortic size and shape into a single measure of morphology, incorporating information from each individual distribution. After training drift-diffusion models on 80% of the available data for each sex, we calculated the square of the Mahalanobis distance for the held-out 20% of data and compared these values to the expected *χ*^2^ distribution. Histograms of the hold-out distances calculated for all 5 folds are shown in Fig. 4. The distribution of hold-out distances matches the *χ*^2^(2) distribution well, with 95% and 97.2% of hold-out data lying within the 95% confidence threshold of the expected distribution for females and males, respectively. This demonstrates the ability of our stochastic models to generalize to unseen normal aortic data.

### 2. Longitudinal validation

The models shown in Fig. 3 were fit on cross-sectional data and evolve from an initial developmental variability of morphology (accounted for by *t*_0_) under the compounding effects of constant drift and diffusion. While each patient contributed a single scan to the model fitting process, a subset of patients also received a second follow-up scan (*n* = 77). As the follow-up data was heldout during fitting, it was used to test the longitudinal fidelity of our stochastic models and determine whether the trajectories of normal aortas evolve as predicted by the model parameters. Fig. 9 displays the trajectories of patient data overlaid on the reference population models in absolute coordinates, and Fig. 10 displays the histogram of squared Mahalanobis distances for the heldout follow-up scans, stratified by sex. The distribution of longitudinal measurements validates our drift-diffusion models, with 93% and 96% of hold-out data lying within the 95% confidence threshold of the expected distribution for females and males, respectively.

**FIG. 9.**
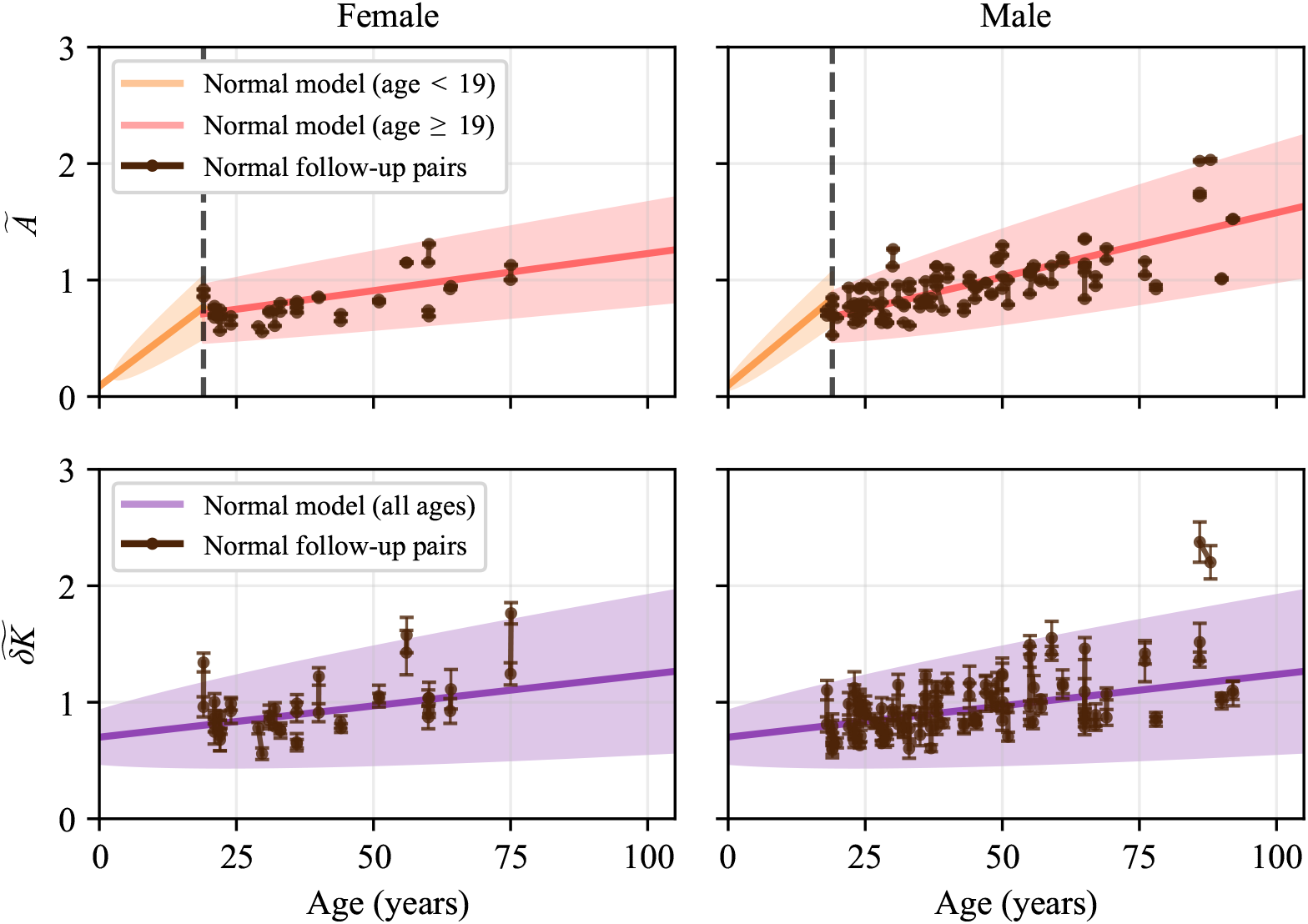
Trajectories of normalized morphologic measurements for non-pathologic patients with multiple scans, with the drift-diffusion models and 95% confidence interval plotted for reference. Each model was fit on single timepoint data that did not include the plotted trajectory measurements. The models’ confidence intervals capture nearly all of the trajectories, indicating their ability to describe normal aortic morphology across repeated measurements. The models demonstrate robustness to random walks in the morphologic space, measurement error, and a range of choices for the discretization of multiple aortic meshes.

**FIG. 10.**
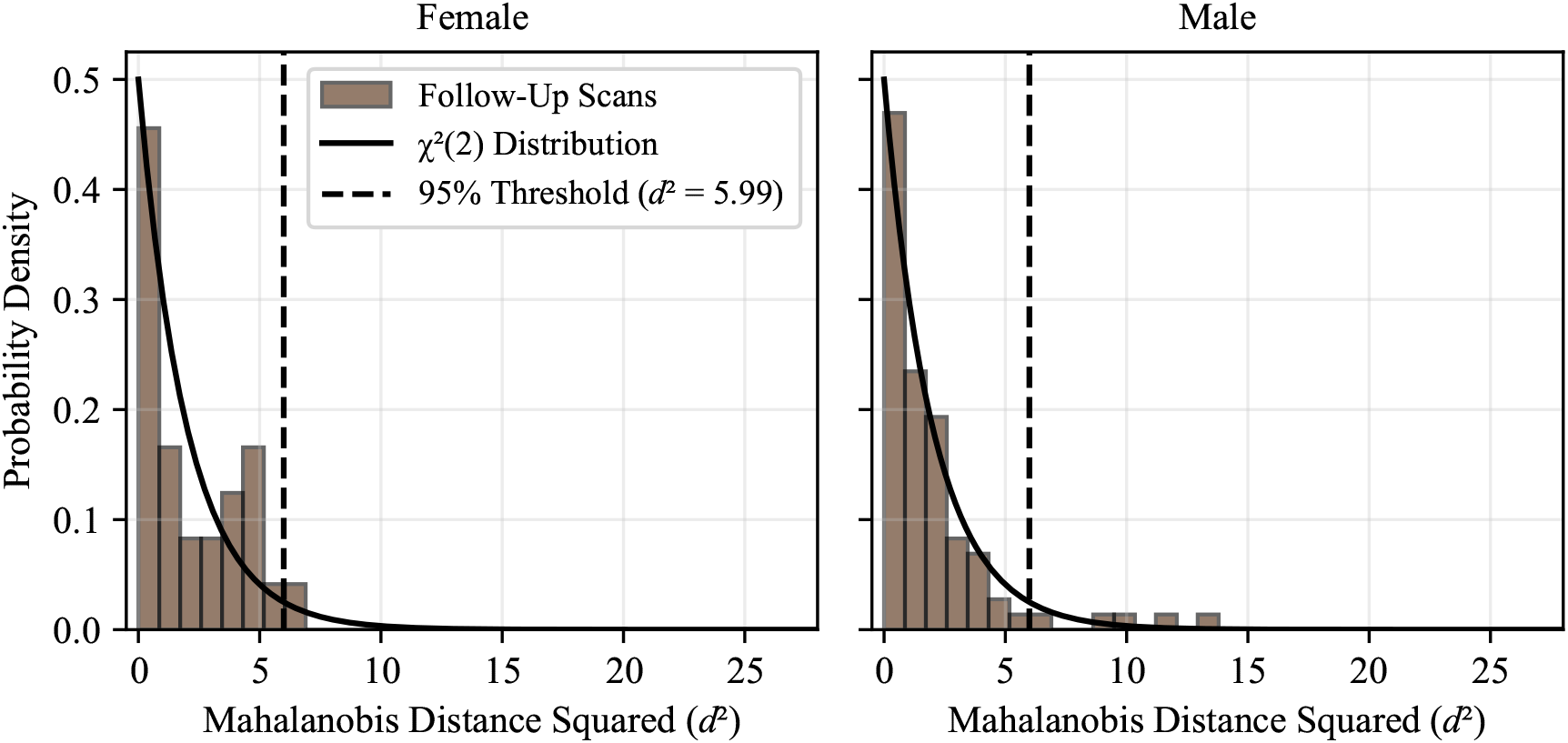
Sex-stratified distributions of *d*^2^ values calculated for a hold-out set of follow-up normal aortic morphologic measurements 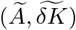 for patients with multiple scans. The drift-diffusion model was fit on all first available normal aortic scans, and the *d*^2^ values for the remaining follow-up data were calculated. The distribution of the hold-out set *d*^2^ values for each sex matches the expected *χ*^2^ distribution well, with 93% and 96% of hold-out data lying within the 95% confidence threshold of the expected distribution for females and males, respectively. This demonstrates the ability of our stochastic models to generalize to unseen, repeated measurements of normal aortic data and accurately describe trajectories of normal aortic morphology.

Because the longitudinal trajectories are not temporally uniform, we also inspect the data on a relative time scale (Fig. 11). With the trajectory data represented as the change in morphology with propagated errors, most of the mean values lie within the corresponding model’s confidence interval (95% of female and 79% of male *Ã* trajectories; 85% of female and 88% of male 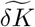 trajectories). Again, this supports the longitudinal validity of the fitted models over the time scale defined by the available data. The relative coordinate space also reveals that of the trajectories that are outside the models’ confidence bounds, more than 80% occur over inter-scan intervals of less than ≈1 month. At these short intervals, the observed morphologic change is dominated by noise inherent to CT-derived surface measurements rather than biological change. Some of this short-interval variability can be attributed to differences in CT acquisition parameters between scans, particularly axial resolution, which affects downstream segmentation, surface mesh quality, and morphology quantification (Figs. 12, 13, 14). Therefore, for longitudinal applications of the framework, an inter-scan interval of at least 1 month is suggested to avoid noise amplification in finite-difference estimates of morphologic change. In a clinical setting, this statistically derived cutoff in 2D morphological space can inform a minimum time frame needed to capture meaningful anatomic changes with CT.

**FIG. 11.**
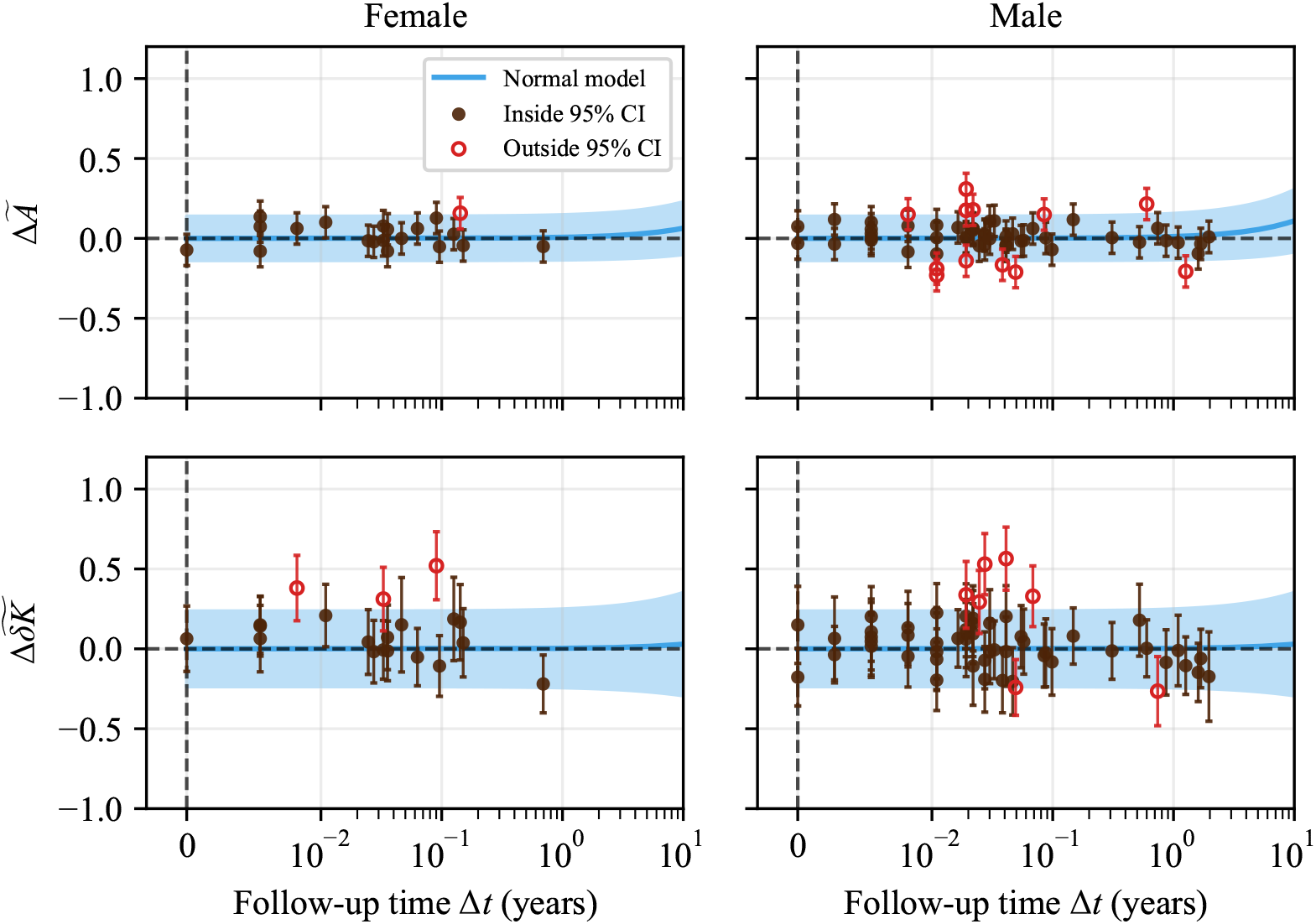
Change in morphology for each non-pathologic patient with two scans, plotted against the time between measurements. For the previously identified adult model parameters, the normal model was propagated from the origin, with the initial 95% confidence interval being calculated from the standard deviation of the difference between measurements for multiple-scan patients with at least 30 days between scans. Of the 23 mean values that lie outside of the 95% confidence interval, 19 (83%) were observed on a time scale of less than 0.1 years. On this short interval, there is inherent noise in test-retest measurements of CT-derived morphology, and for studies involving the time derivative of morphologic change, this approximate time scale can serve as a boundary for the minimum time that needs to pass between scans to remain confident that unnecessary noise amplification is avoided in finite-difference calculation.

**FIG. 12.**
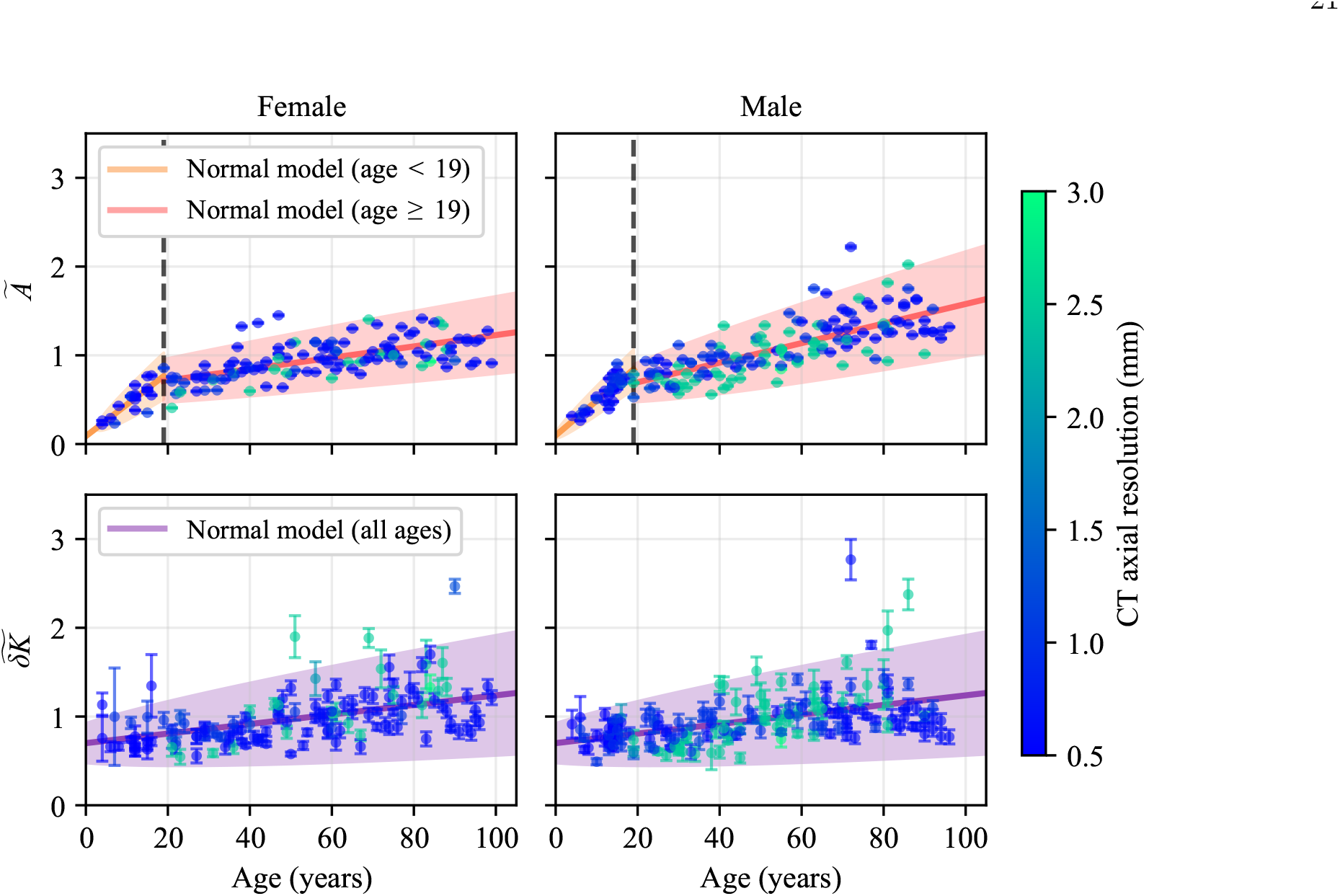
Normalized aortic surface area (*Ã*) and fluctuation in integrated Gaussian curvature 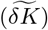 measurements colored by each scan’s axial resolution. Models fit on the full dataset (all resolutions) underlie the raw points.

**FIG. 13.**
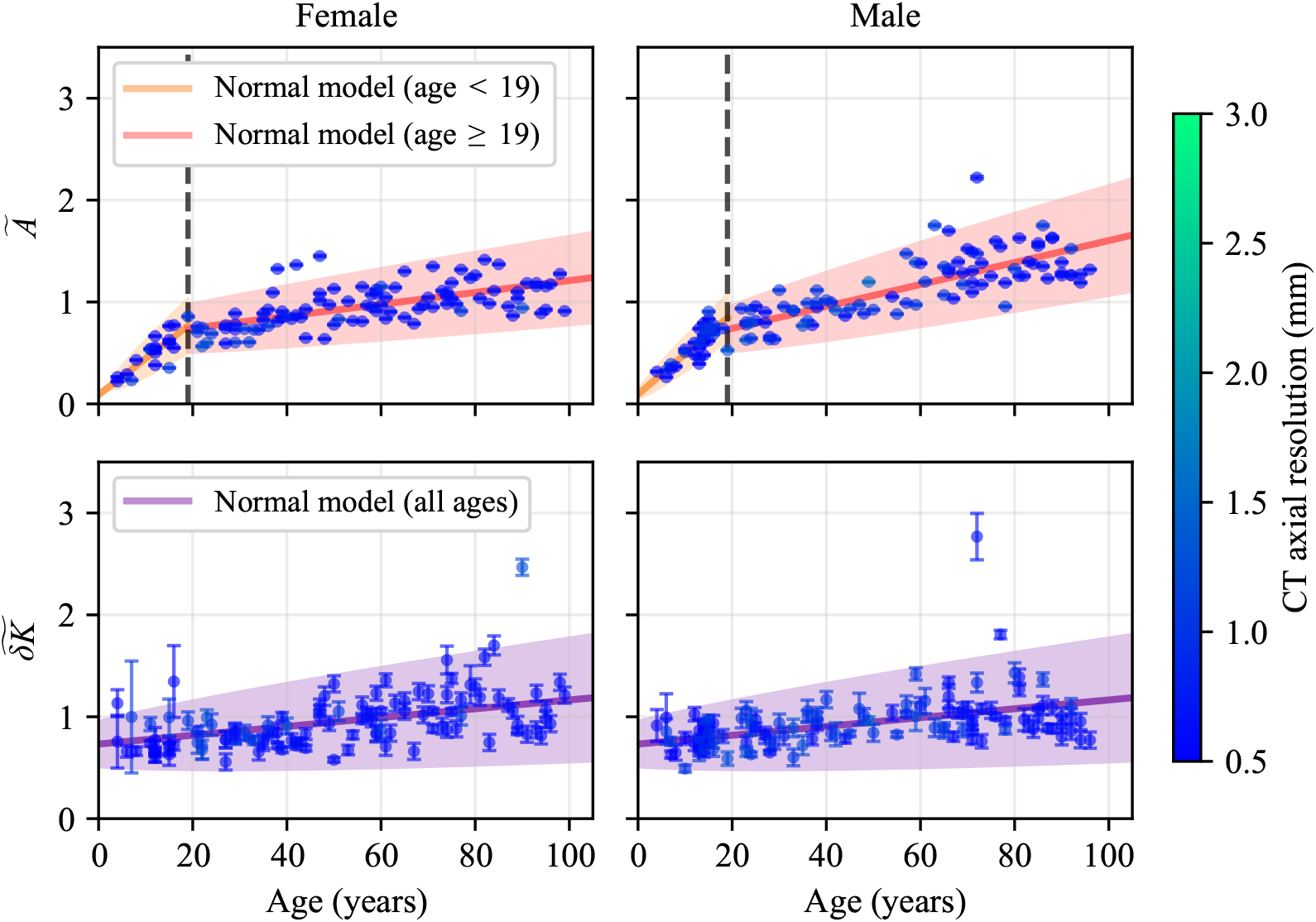
Subset of data with axial resolution ≤1.5 mm, with models re-fit on this subset. Comparison with Fig. 12 confirms that the fitted mean trajectories and 95% confidence envelopes are minimally affected by restriction to high-resolution scans. Per-parameter changes are reported in Table VII.

**FIG. 14.**
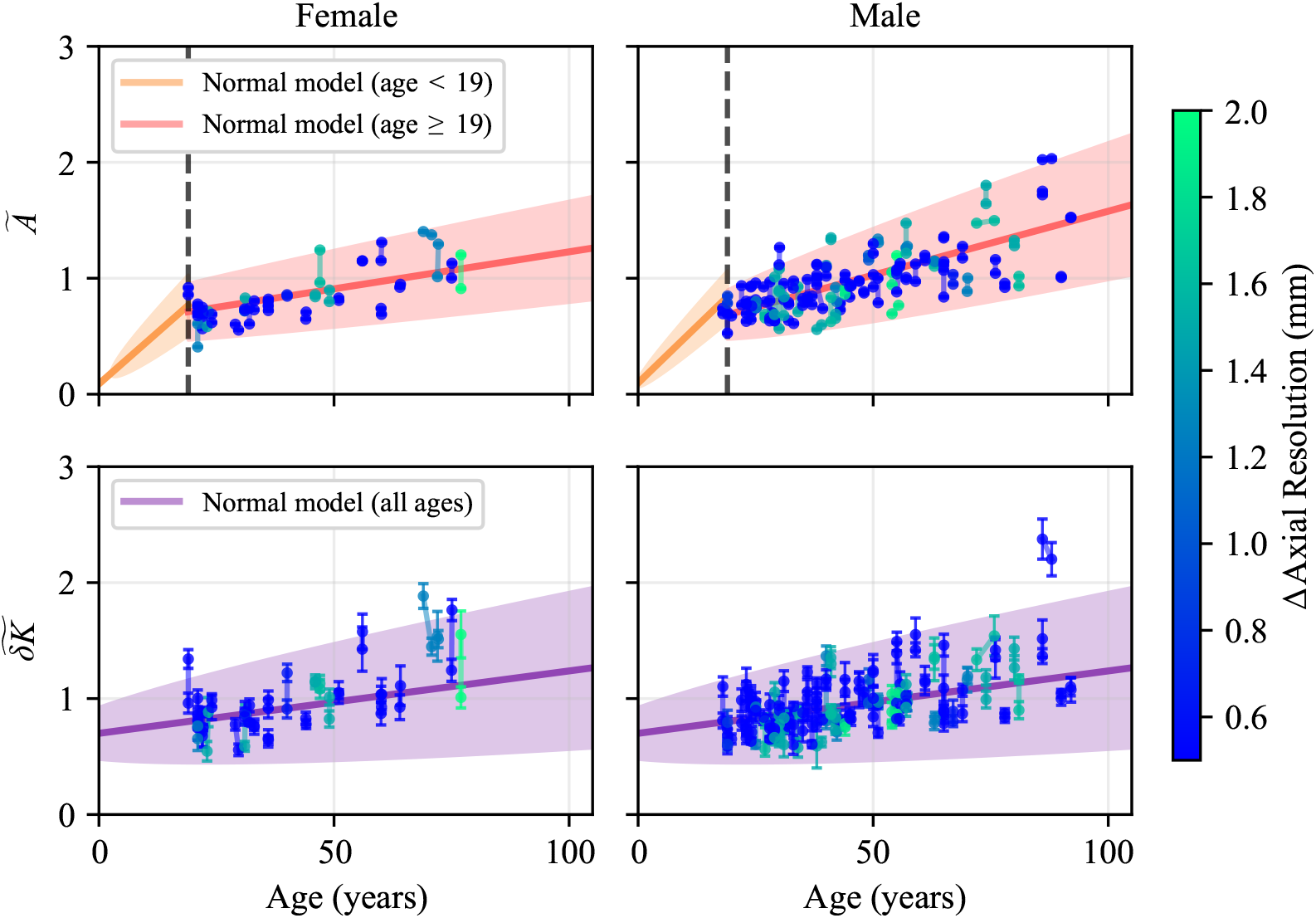
Trajectories of *Ã* and 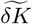 for multi-scan patients, colored by the absolute difference in axial resolution between scans. Trajectories with large apparent morphologic change between scans are concentrated in pairs with large resolution change. The 36 patients (31.9%) with between-scan resolution change exceeding 1 mm show significantly larger absolute change in *Ã* (0.120 vs. 0.074, Mann-Whitney U one-sided *p* = 0.013) than the 77 patients with |Δ*z*| *≤* 1 mm.

## Appendix K: Model robustness to variation in CT resolution

Any CT-derived morphologic analysis must contend with variability introduced by imaging acquisition and segmentation [25, 51, 73]. Because this is a retrospective study of clinically acquired data, our dataset consists of scans taken by multiple machines, under distinct acquisition parameters, by different technicians, and under different clinical orders. We addressed these sources of variability where possible while preserving the heterogeneity inherent to a clinical dataset.

### 1. Cross-sectional modeling with heterogeneous resolutions

We first assess the impact of CT resolution on the cross-sectional model fits by identifying the axial resolution corresponding to each measurement and re-fitting the stochastic models on the subset of data with resolution ≤1.5 mm. Then, we compare both the fitted parameters and the resulting envelopes to those obtained from the full dataset.

Fig. 12 shows the full dataset colored by axial resolution with models fit on all data, while Fig. 13 shows the high-resolution subset with models fit on that subset alone. The fitted mean trajectories and 95% confidence envelopes are visually similar between the two fits across all four panels, demonstrating that the model’s predictions are minimally affected by the inclusion of lower-resolution scans. Per-parameter changes are reported in Table VII: the drift coefficient *β* shifts by less than 10% for both sexes’ *Ã* models, with the female child model unchanged because all pediatric scans were already ≤ 1.5 mm. The diffusion coefficient *α* and the reference time *t*_0_ show larger relative shifts in some models—most notably 20.5% in *α* for adult male *Ã* and 60.1% in *t*_0_— but these reflect tradeoffs along the drift direction (*t*_0_ with *x*_0_) and noise-floor sensitivity (*α*) that do not propagate to the model’s predicted trajectories at observed ages, consistent with the visual equivalence of the envelopes in the two figures. Restricting to ≤ 1.5 mm scans also removes a number of outlier measurements visible in Fig. 12, contributing to the reduction in *α* observed for the sex-aggregated, single-regime 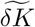 model in Table VII. The qualitative findings—sex-stratified and sex-aggregated growth rates for *Ã* and 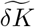 respectively, the developmental-to-adult regime transition for size, and the single-regime structure for shape—are preserved between the two fits, supporting the model’s robustness to CT scan quality and its translatability to clinical settings where acquisition parameters vary.

**TABLE VII.**
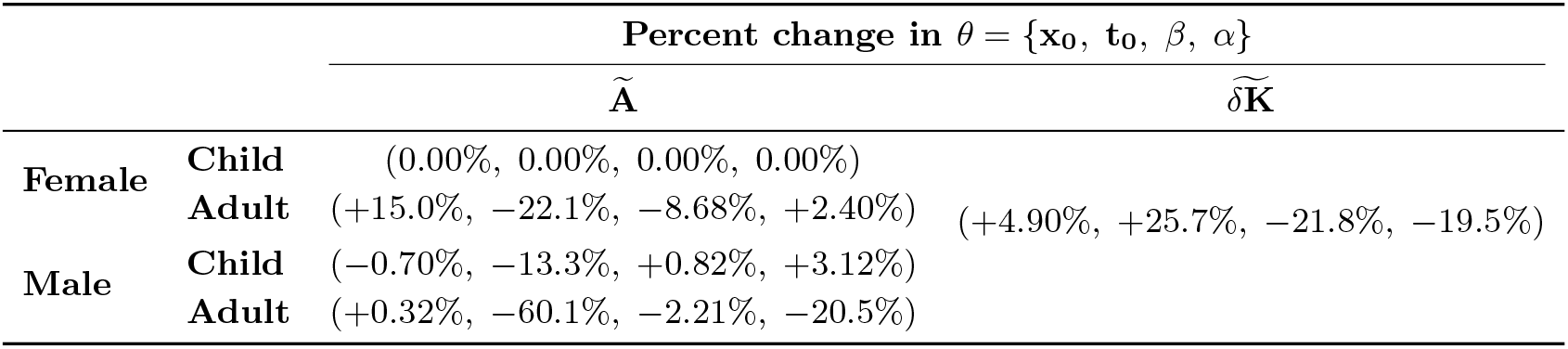
Percent change in fitted parameters between models fit on the full dataset and on the high-resolution subset (*≤* 1.5 mm axial resolution). The female child model of *Ã* is unchanged because all pediatric scans were already ≤ 1.5 mm. Large relative changes in *t*_0_ correspond to small absolute shifts in a parameter that trades off with *x*_0_ along the drift direction, so they do not substantially affect the predicted mean trajectory at observed ages.

### 2. Longitudinal sensitivity to between-scan resolution change

In contrast to cross-sectional model fits, longitudinal analyses of within-patient morphologic change are sensitive to between-scan changes in axial resolution. Fig. 14 displays the trajectories of multi-scan patients colored by the absolute difference in axial resolution between their two scans, where trajectories with the largest apparent morphologic change are concentrated in the pairs with the largest resolution change.

Of the 113 multi-scan patients with paired axial resolution data, 36 (31.9%) had a between-scan resolution change exceeding 1 mm. These patients had a mean absolute change in *Ã* of 0.120 over a mean follow-up of 0.275 years, compared with a mean absolute change of 0.074 for the 77 patients with |Δ*z*|≤ 1 mm (Mann-Whitney U one-sided *p* = 0.013). Although the above-threshold group had a longer mean follow-up time (0.28 vs 0.18 years), the magnitude of the difference in observed *Ã* substantially exceeds what would be expected from biological growth alone over that interval based on the fitted adult drift rates (Table I). The corresponding effect for 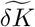 was in the same direction (mean change of 0.174 vs 0.136) but did not reach statistical significance (*p* = 0.16), suggesting either greater inherent robustness of the shape metric to resolution-driven artifacts or insufficient power against the higher baseline noise of the curvature measurement. The magnitude of this resolution-driven effect also substantially exceeds the parameterization-dependent error quantified by the scale-space framework of Pugar et al. [51]. We therefore restricted the longitudinal analysis in the main text (Fig. 9) to trajectories with a between-scan resolution change of *<* 1 mm. The residual variability after this filter reflects the inherent error of segmentation, which is unavoidable across studies analyzing segmented surfaces. Standardization of CT acquisition protocols, particularly consistent axial slice thickness across serial imaging, would further improve the reliability of longitudinal morphologic assessment and is an important consideration for prospective study design.

